# Adherence to a healthful plant-based diet and risk of mortality among individuals with chronic kidney disease: A prospective cohort study

**DOI:** 10.1101/2024.04.08.24305486

**Authors:** Alysha S. Thompson, Martina Gaggl, Nicola P. Bondonno, Amy Jennings, Joshua K. O’Neill, Claire Hill, Nena Karavasiloglou, Sabine Rohrmann, Aedín Cassidy, Tilman Kühn

**Author notes:** **Correspondence:** and **Corresponding Authors:** Prof Dr Tilman Kuhn and Prof Aedín Cassidy Queen’s University Belfast Institute for Global Food Security (IGFS) / School of Biological Sciences 19 Chlorine Gardens Belfast BT9 5DL UK. **Funding Source:** Alysha S. Thompson holds a PhD studentship of the Department for the Economy (DfE), Northern Ireland.

## Abstract

**Background:** Plant-rich dietary patterns may protect against negative health outcomes among individuals with chronic kidney disease (CKD), although aspects of plant-based diet quality have not been studied. This study aimed to examine associations between healthful and unhealthful plant-based dietary patterns with risk of mortality among CKD patients for the first time.

**Methods:** This prospective analysis included 4,807 UK Biobank participants with CKD at baseline. We examined associations of adherence to both the healthful plant-based diet index (hPDI) and unhealthful plant-based diet index (uPDI), calculated from repeated 24-hour dietary assessments, with risk of mortality using multivariable Cox proportional hazard regression models.

**Results:** Over a 10-year follow-up, 675 deaths were recorded. Participants with the highest hPDI scores had a 33% lower risk of mortality [HR_Q4vsQ1_ (95% CI): 0.67 (0.53-0.84), p_trend_ = <0.001], while those with the highest uPDI scores had a 49% higher risk [1.49 (1.18-1.89), p_trend_ = 0.004], compared to participants with the lowest respective scores and following adjustment for other dietary and lifestyle factors. In food group-specific analyses, higher wholegrain intakes were associated with a 29% lower mortality risk, while intakes of refined grains, and sugar-sweetened beverages were associated a 28% and 31% higher risk, respectively.

**Conclusions:** In CKD patients, a higher intake of healthy plant-based foods was associated with a lower risk of mortality, while a higher intake of unhealthy plant-based foods was associated with a higher risk. These results underscore the importance of plant food quality and support the potential role of healthy plant food consumption in the treatment and management of CKD to mitigate unfavourable outcomes.

## 1. Introduction

Chronic kidney disease (CKD) poses a significant global public health challenge. The number of CKD-attributed deaths has increased by 98% from 1990 to 2016 (∼0.6 million to ∼1.2 million deaths), with projections that CKD will rank fifth among the leading causes of death by 2040 (1–3). Further, the burden of disability-adjusted life-years (DALYs) attributed to CKD has risen from 29^th^ in 1990 to the 18^th^ in 2019 (4). CKD is a progressive and comorbid condition, often leading to end-stage kidney disease (ESKD), necessitating kidney replacement therapy such as dialysis or transplantation (3, 5, 6). Notably, CKD stands out as one of the few non-communicable diseases to sustain a continual increase in associated mortality over the past two decades (1). Given CKD’s substantial impact on the economy (7) and on quality of life (8), there is a pressing need to explore how dietary and lifestyle modifications can preserve kidney function and improve outcomes in CKD patients.

A plant-based diet (PBD) including lower amounts of animal-based foods has previously been shown to be associated with better outcomes e.g., improved insulin sensitivity and lower blood pressure and survival in individuals living with CKD (9–12). Previously, a diet promoting solely plant proteins was criticised for enhancing protein-energy wasting and malnutrition, especially in patients with CKD stage 5, however, more recent studies suggest that plant-based proteins are both nutritionally adequate and beneficial for CKD patients (13). Further, vegetarian, and vegan diets are now increasingly used to facilitate better adherence to the 2020 Kidney Disease Outcomes Quality Initiative (KDOQI) nutrition guideline which advises patients with earlier stages of CKD to lower dietary protein intake to mitigate further kidney damage caused by glomerular hyperfiltration (14, 15).

While dietary modification to treat and manage CKD has a long tradition (16–18), the relationship between PBD quality in the form of healthful and unhealthful plant-based dietary patterns with risk of mortality in patients with early-stage CKD is unknown. Therefore, this study aims to investigate the relationship between plant-based diet indices (PDIs) and the risk of all-cause mortality among individuals with CKD using data from the UK Biobank cohort.

## 2. Methods

### 2.1 Study population

The UK Biobank is a large cohort study of over 500,000 middle-aged adults recruited between 2006-2010 from the United Kingdom. Participants were recruited across 22 study centres in England, Scotland and Wales and provided baseline data by completing touchscreen questionnaires, verbal interviews and undergoing physical and biological assessments (19). The UK Biobank was approved by the NHS North West Multicentre Research Ethics Committee (Ref. 11/NW/0382). All study participants provided written informed consent at recruitment.

For this study, 502,236 participants were screened for a prior diagnosis of CKD and available data from at least one valid 24-hr dietary assessment. Participants who withdrew their consent during follow-up, had implausible energy intakes (>17,573KJ or < 3,347KJ for men and > 14,644KJ or < 2,092KJ for women (20)), had missing covariate information, or their follow-up ended before completing the final 24-hr dietary assessment were excluded from this study. Of the 31,556 UK Biobank participants with CKD, 4,807 completed at least 1 dietary assessment and were included in the present analyses (**Supplementary Figure S1).**

### 2.2 Dietary assessment

Dietary intake was measured using the Oxford WebQ, a self-administered and internet-based 24-hr dietary assessment tool. The Oxford WebQ assessment was issued on five separate occasions: once during the recruitment phase as part of the baseline assessment at the study centre (April 2009 – September 2010), and then on four further instances via home computer outside of the recruitment period (February 2011 – June 2012). Between April 2009 and June 2012, a minimum of 1 and a maximum of 5 assessments were completed. Mean (SD) duration (years) between baseline (cycle 0) and first (cycle 1) and last (cycle 4) dietary assessments were 1.6 (1.4) and 2.3 (1.4), respectively. Further details on the Oxford WebQ have been documented elsewhere (21). The reliability of the Oxford WebQ is supported by validation studies (22–24).

### 2.3 Plant-based diet indices

Following methods described previously (25–27), the Oxford WebQ tool was used to construct a healthful plant-based diet index (hPDI) and unhealthful plant-based diet index (uPDI); an established scoring tool used to assess the overall adherence to a PBD, considering the quality of plant food consumption. To calculate the indices, 17 food groups were used (whole grains, fruits, vegetables, nuts, legumes and vegetarian protein alternatives, tea and coffee, fruit juices, refined grains, potatoes, sugar-sweetened beverages (SSBs), sweets and desserts, animal fat, dairy, eggs, fish or seafood, meat, and miscellaneous animal-based foods). Unlike previous studies where vegetable oils were included as a ‘healthy’ food component, data on vegetable oils was not available for this study. For the hPDI, healthy plant-food groups were scored positively, and unhealthy plant-food groups and animal-derived food groups were scored in the reverse. For the uPDI, positive scores were assigned to unhealthy plant-food groups and reverse scores were given to healthy plant-food groups and animal-derived food groups. For each food group, intakes of more than zero servings were ranked into sex-specific quartiles. For positive scores, participants were assigned a score between 2 and 5 (2 for the lowest quartile of intake and 5 for the highest). Participants who reported zero intakes were assigned a score of 1. The pattern of scoring was inverted for reverse scores. The scores assigned to each of the 17 food groups were summed to obtain the final hPDI and uPDI.

### 2.4 Ascertainment of chronic kidney disease

CKD cases were considered prevalent if diagnosed before the first dietary assessment via self-reported or hospital inpatient data. CKD cases were defined using the International Classification of Diseases, 10 Revision (ICD-10) codes (ICD-10 codes: N03, N06, N08, N11, N12, N13, N14, N15, N16, N18, N19, Z49, I12, I13) or Office of Population Censuses and Surveys Classification of Interventions and Procedures-version 4 (OPCS-4) codes in Hospital Inpatient data (OPCS-4 codes: L74.1-L74.6, L74.8, L74.9, M01.2, M01.3-M01.5, M01.8, M01.9, M02.3, M08.4, M17.2, M17.4, M17.8, M17.9, X40.1-X40.9, X41.1, X41.2, X41.8, X41.9, X42.1, X42.8, X42.9, and X43.1). Further information regarding the codes used to define CKD can be found in **Supplement Table S1** . Using serum creatinine, the CKD-EPI (Chronic Kidney Disease Epidemiology Collaboration) creatinine equation (28) was used to calculate glomerular filtration rate. Participants with a baseline estimated glomerular filtration rate (eGFR) <60ml/min/1.73m were considered as having CKD.

### 2.5 Ascertainment of mortality

The primary outcome of this study was all-cause mortality. Death data was obtained from death certificates provided by the National Health Service England (England and Wales) and National Health Service Central Register, National Records of Scotland (Scotland). Mortality data was available until 30 November 2022 for England, Scotland, and Wales. Follow-up time was calculated from the date of the last dietary assessment and was censored at date of death, lost to follow-up, or death registry censoring date, whichever was the earliest.

### 2.6 Assessment of covariates

Between 2006 and 2010, participants provided information on sociodemographic, dietary, lifestyle and medical history at the assessment centre, where several types of assessments were performed. A touchscreen questionnaire was used to collect information on self-reported age, sex, ethnicity, smoking status, education level and physical activity at baseline. Multimorbidity was identified through data on past and current medical conditions obtained through verbal interview at the initial assessment. Polypharmacy was also obtained through verbal interview at baseline and was defined by the number of self-reported treatments/medications taken. Prevalent diabetes was defined through self-reporting at the touchscreen questionnaire and verbal interview. Townsend deprivation index was calculated at baseline using postcodes of residence. Physical measures including waist circumference, height, and weight were manually obtained at the initial assessment by trained professionals. Height and weight were used to calculate body mass index (BMI), dividing weight (kg) by the square of height (m). Number of completed dietary assessments, energy, alcohol, and protein intake was based on the 24-hr Oxford WebQ dietary questionnaire. To calculate eGFR, the CKD-EPI (Chronic Kidney Disease Epidemiology Collaboration) creatinine equation (28) was employed using serum creatinine which was measured by enzymatic analysis on a Beckman Coulter AU5800.

### 2.7 Assessment of genetic risk score of eGFR

A detailed description of the genotyping, imputation, and quality control of the UK Biobank genetic data has been published previously (29). A recent GWAS by Wuttke et al. 2019 was used to identify single nucleotide polymorphisms (SNPs) associated with CKD. Using a weighted method, a polygenic risk score (PRS) of eGFR was calculated using 161 SNPs (**Supplementary Table S2** ). A higher PRS of eGFR indicates a lower genetic risk of kidney diseases. Additional information on the quality control procedures implemented in this study have been described in the **Supplementary Methods**.

### 2.8 Statistical analysis

Baseline characteristics of the study population are presented across quartiles of hPDI and uPDI scores. Continuous variables are presented as mean ± standard deviation (SD) and categorical variables are presented as a number and percentage proportion. Associations between PDIs and total mortality were examined using Cox proportional hazards regression models, with age (years) as the timescale. Results are reported as hazard ratios (HRs) and 95% confidence intervals (95% CI). Linear trend tests involved modelling the PDIs as continuous exposure variables (P_trend_). The proportional hazards assumption was tested by the Schoenfeld residual method. Analyses were adjusted for confounding factors using two models. Model 1 was adjusted for sex (female, male) and education (Low: CSEs or equivalent, O levels/GCSEs or equivalent; Medium: A levels/AS levels or equivalent, NVQ or HND or HNC or equivalent; High: College or University degree, other professional qualifications eg: nursing, teaching; unknown/missing/prefer not to say (15.6%)), stratified by age (5-year categories) and geographical region of recruitment (ten UK regions). Model 2 was additionally adjusted for BMI (18·5-24·9 kg/m^2^, 25·0-29·9 kg/m^2^, ≥30 kg/m^2^, or unknown/missing (0.4%)), waist circumference (continuous scale, cm), ethnicity (Asian, Black, Multiple, White, other/unknown/missing (1.3%)), physical activity (Metabolic Equivalent Tasks (METs) hr/week in quintiles, or unknown/missing (2.7%)), smoking status (never, previous, current, or unknown/missing (0.3%)), alcohol intake (continuous scale, g/day), energy intake (continuous scale, kJ/day), multimorbidity index (number of pre-existing long-term conditions at baseline; 0, 1, 2, or >3), polypharmacy index (total number of self-reported medications take at baseline; 0, 1-3, 4-6, 7-9, >10), Townsend deprivation index (quintiles from low to high deprivation), prevalent diabetes (no, yes), protein intake (continuous, g/day) and number of completed dietary assessments (continuous scale, ranging between 1-5). In addition to main analyses, multivariable Cox regression analyses were performed for each of the 17 PDI food groups.

To investigate whether associations between the continuous hPDI and uPDI (10-point increments) and risk of mortality differed by population groups, analyses were stratified by age (<63 or ≥63 years), sex (male or female), smoking status (never or ever), alcohol intake (tertiles from low to high alcohol intake: low [<1g/day], moderate [1-≤14g/day] or high [≥15g/day]), protein intake (<median [<77g/day] or ≥ median [≥77g/day]), eGFR (<35 mL/min/1,73 m^2^ or ≥35 mL/min/1,73 m^2^ ), PRS (tertiles from low to high PRS for eGFR) and CKD stage (stage 1-2, stage 3a-3b, or stage 4-5), testing for interactions using likelihood ratio tests (LRT). To assess the robustness of our findings, three further sets of analyses were conducted. Firstly, primary analyses were further adjusted for eGFR to account for level of kidney function. Secondly, primary analyses were further adjusted for genetic susceptibility to kidney diseases (PRS of eGFR). Thirdly, analyses were restricted to participants who had completed ≥2 dietary assessments. Finally, to assess potential reverse causation bias, sensitivity analyses were carried out excluding the first 2 years of follow-up.

Intraclass correlation coefficients (ICCs) were used to test the reliability of the PDIs over time. PDI scores were calculated from mean food intakes at each Oxford WebQ dietary assessment (cycle 0: April 2009 to September 2010 vs cycle 1: February 2011 to April 2011 vs cycle 2: June 2011 to September 2011 vs cycle 3: October 2011 to December 2011 vs cycle 4: April 2012 to June 2012).

Statistical analyses were performed using Stata statistical software version 18.0 (StataCorp LLC). Two-sided P values were considered statistically significant.

## 3. Results

### 3.1 Study participants and baseline characteristics

Baseline characteristics and nutrient intakes of 4,807 study participants with CKD are shown across quartiles of hPDI and uPDI in **Table 1 and Supplementary Table S3-Table S5.** A comparison of baseline characteristics of CKD patients with and without dietary data is shown in **Supplementary Table S6** . The mean (SD) age was 60.9 (6.9), 2,452 (51.0%) were female, 543 (11.3%) had prevalent diabetes, and the mean baseline eGFR was 59.5 mL/min/1.73m^2^. Individuals with higher scores of hPDI were more likely to be women, have a lower BMI, be more physically active and have a higher education level than those with lower scores. Individuals with higher scores of uPDI were more likely to be men, have a higher BMI, be less physically active and have a lower education level than those with lower scores.

**Table 1.** Baseline characteristics across quartiles (Q) of the healthful plant-based diet index in the UK Biobank (n=4,807)

| Characteristics across hPDI | Participants, No. (%)<sup>a</sup> |  |  |  |  |
| --- | --- | --- | --- | --- | --- |
|  | Q1 | Q2 | Q3 | Q4 | Whole sample |
| **Number of participants** | 1,309 (27.2) | 1,239 (25.8) | 1,131 (25.5) | 1,128 (23.5) | 4,807 (100.0) |
| **Mortality cases** | 236 (18.0) | 168 (13.6) | 140 (12.4) | 131 (11.6) | 675 (14.0) |
| **Healthful plant-based diet index, mean (SD)** | 49.7 (3.2) | 55.6 (1.5) | 59.5 (1.5) | 64.9 (2.7) | 57.1 (6.0) |
| **Sex-Female** | 631 (48.2) | 631 (50.9) | 598 (52.9) | 592 (52.5) | 2,452 (51.0) |
| **Age at recruitment (years), mean (SD)** | 60.1 (7.3) | 61.1 (6.9) | 61.5 (6.6) | 61.3 (6.5) | 60.9 (6.5) |
| **BMI (kg/m<sup>2</sup>), mean (SD)** | 29.5 (5.8) | 28.5 (4.9) | 28.2 (5.0) | 27.7 (4.7) | 28.5 (5.2) |
| **Waist circumference (cm), mean (SD)** | 96.9 (14.9) | 93.9 (14.0) | 93.1 (13.5) | 91.9 (13.3) | 94.0 (14.1) |
| **Energy intake (kJ/day), mean (SD)** | 9125.7 (2298.2) | 8370.9 (2107.8) | 7997.0 (2139.7) | 7810.8 (2118.5) | 8357.1 (2229.9) |
| **Physical activity (MET-h/wk), mean (SD)** | 28.6 (45.8) | 28.2 (38.3) | 29.4 (36.0) | 33.8 (43.2) | 29.9 (41.2) |
| **Ethnicity** |  |  |  |  |  |
| Asian | 47 (3.6) | 46 (3.7) | 43 (3.8) | 37 (3.3) | 173 (3.6) |
| Black | 1 (0.1) | 3 (0.2) | 7 (0.6) | 2 (0.2) | 13 (0.3) |
| Multiple | 41 (3.1) | 46 (3.7) | 32 (2.8) | 39 (3.5) | 158 (3.3) |
| White | 1,205 (92.1) | 1,130 (91.2) | 1,034 (91.4) | 1,033 (91.6) | 4,402 (91.6) |
| Other/missing<sup>b</sup> | 15 (1.2) | 14 (1.1) | 15 (1.3) | 17 (1.5) | 61 (1.3) |
| **Education<sup>c</sup>** |  |  |  |  |  |
| Low | 332 (25.4) | 282 (22.8) | 280 (24.8) | 257 (22.8) | 1,152 (24.0) |
| Medium | 211 (16.1) | 200 (16.1) | 177 (15.7) | 199 (17.6) | 787 (16.4) |
| High | 522 (40.0) | 548 (44.2) | 515 (45.5) | 531 (47.1) | 2,116 (44.0) |
| Missing | 244 (18.6) | 208 (16.8) | 159 (14.1) | 141 (12.5) | 752 (15.6) |
| **Smoking status** |  |  |  |  |  |
| Never | 670 (51.2) | 666 (53.8) | 571 (50.5) | 597 (52.9) | 2,504 (52.1) |
| Previous | 527 (40.3) | 497 (40.1) | 500 (44.2) | 466 (41.3) | 1,990 (41.4) |
| Current | 110 (8.4) | 71 (5.7) | 56 (5.0) | 62 (5.5) | 299 (6.2) |
| Missing | 2 (0.2) | 5 (0.4) | 4 (0.6) | 3 (0.3) | 14 (0.3) |
| **Alcohol intake (g/day), mean (SD)** | 13.4 (19.6) | 13.5 (18.5) | 13.6 (19.4) | 12.3 (19.6) | 13.2 (19.3) |
| **Prevalent diabetes** | 168 (12.8) | 136 (11.0) | 112 (9.9) | 127 (11.3) | 543 (11.3) |
| **Multimorbidity** |  |  |  |  |  |
| 0 LTCs | 361 (27.6) | 343 (27.7) | 333 (29.4) | 334 (29.6) | 1,371 (28.5) |
| 1 LTC | 315 (24.1) | 317 (25.6) | 317 (28.0) | 293 (26.0) | 1,242 (25.8) |
| 2 LTCs | 302 (23.1) | 296 (23.9) | 247 (21.8) | 275 (24.4) | 1,120 (23.3) |
| ≥3 LTCs | 331 (25.3) | 283 (22.8) | 234 (20.7) | 226 (20.0) | 1,074 (22.3) |
| **eGFR (mL/min/1.73 m<sup>2</sup>), mean (SD)** | 58.9 (18.6) | 59.2 (18.0) | 58.7 (16.6) | 61.3 (18.4) | 59.5 (18.0) |
| **Protein intake (g/day), mean (SD)** | 83.9 (25.0) | 79.6 (22.6) | 77.7 (23.6) | 76.9 (22.4) | 79.7 (23.6) |
| **hPDI food item intake (portion/day)<sup>d</sup>, mean (SD)** |  |  |  |  |  |
| **Healthy plant food** |  |  |  |  |  |
| Whole grains | 1.4 (1.4) | 1.9 (1.6) | 2.2 (1.6) | 2.7 (1.7) | 2.0 (1.6) |
| Fruit | 1.5 (1.3) | 1.9 (1.5) | 2.3 (1.7) | 3.1 (1.9) | 2.2 (1.7) |
| Vegetables | 1.6 (1.6) | 2.0 (1.9) | 2.5 (2.2) | 3.4 (2.5) | 2.4 (2.2) |
| Nuts | 0.1 (0.2) | 0.1 (0.3) | 0.1 (0.3) | 0.2 (0.5) | 0.1 (0.4) |
| Legumes | 0.2 (0.4) | 0.3 (0.5) | 0.4 (0.5) | 0.6 (0.7) | 0.4 (0.5) |
| Tea and coffee | 3.8 (1.8) | 4.1 (1.7) | 4.5 (1.8) | 4.8 (1.8) | 4.3 (1.8) |
| **Unhealthy plant food** |  |  |  |  |  |
| Refined grains | 1.9 (1.5) | 1.2 (1.2) | 0.9 (1.1) | 0.6 (0.9) | 1.2 (1.3) |
| Potatoes | 0.9 (0.7) | 0.8 (0.7) | 0.7 (0.6) | 0.6 (0.6) | 0.7 (0.6) |
| Sugar-sweetened beverages | 1.0 (1.2) | 0.6 (0.9) | 0.4 (0.8) | 0.3 (0.7) | 0.6 (1.0) |
| Fruit juices | 0.5 (0.6) | 0.5 (0.6) | 0.4 (0.6) | 0.3 (0.6) | 0.4 (0.6) |
| Sweets and desserts | 2.0 (1.6) | 1.6 (1.3) | 1.3 (1.3) | 0.9 (1.1) | 1.5 (1.4) |
| **Animal-derived food** |  |  |  |  |  |
| Animal fat | 1.3 (1.4) | 0.8 (1.2) | 0.5 (1.0) | 0.3 (0.7) | 0.8 (1.2) |
| Dairy | 1.2 (0.9) | 1.1 (0.8) | 1.0 (0.8) | 0.9 (0.8) | 1.0 (0.9) |
| Eggs | 0.4 (0.6) | 0.3 (0.5) | 0.2 (0.5) | 0.2 (0.4) | 0.3 (0.5) |
| Fish or seafood | 0.4 (0.5) | 0.3 (0.5) | 0.3 (0.5) | 0.2 (0.4) | 0.3 (0.5) |
| Meat | 1.5 (1.1) | 1.3 (1.0) | 1.1 (0.9) | 1.0 (0.8) | 1.2 (1.0) |
| Miscellaneous animal-based foods | 0.1 (0.4) | 0.1 (0.2) | 0.1 (0.3) | 0.0 (0.2) | 0.1 (0.3) |

<sup>a</sup>Relative frequencies (%) include missing values which may not equate to 100%.
<sup>b</sup>Other includes any race or ethnic group not otherwise specified.
<sup>c</sup>Education was categorised as Low: CSEs or equivalent, O levels/GCSEs or equivalent; Medium: A levels/AS levels or equivalent, NVQ or HND or HNC or equivalent; High: College or University degree, other professional qualifications eg: nursing, teaching.
<sup>d</sup>Portion sizes were specified as a “serving” in the Oxford WebQ tool.
*Abbreviations: Q, quartile; hPDI, healthful plant-based diet index; BMI, body mass index; MET, metabolic equivalent task; SD, standard deviation; LTC, long term condition; eGFR, estimated glomerular filtration rate.*

The ICCs (range) for reproducibility of the PDIs over time ranged from 0.48 (39–75) to 0.53 (45–79) for hPDI and 0.42 (42–72) to 0.48 (40–73) for uPDI, respectively (**Supplementary Table S7)**.

### 3.2 Plant-based diets and mortality risk

During a mean (IQR) follow-up of 10.4 (0.9) years, 675 participants died. In multivariable-adjusted Cox regression models (Model 2), dose-response associations were shown for both hPDI and uPDI with risk of mortality. Participants in the highest hPDI quartile (Quartile 4 (Q4)) had a 33% lower risk of mortality [HR _Q4vsQ1_ (95% CI): 0.67 (0.53-0.84); p_trend_ = 0.001] compared to those in the lowest hPDI quartile (Quartile 1 (Q1)) (**Table 2** ). In contrast, participants in the highest uPDI quartile (Q4) had 49% higher risk of mortality [1.49 (1.18-1.89); p_trend_ = 0.004] compared to those in the lowest uPDI quartile (Q1).

**Table 2.** Hazard ratios (95% confidence intervals) of mortality among all individuals with chronic kidney disease across sex-specific quartiles (Q) of the healthful plant-based diet index (hPDI) and unhealthful plant-based diet index (uPDI) (n=4,807)

| **hPDI** | **Q1** | **Q2** | **Q3** | **Q4** | **P-trend** |
| --- | --- | --- | --- | --- | --- |
| Cases/total | 236/1,309 | 168/1,239 | 140/1,131 | 131/1,128 |  |
| HR (95% CI) |  |  |  |  |  |
| Model 1 | 1.00<sup>a</sup> | 0.71 (0.58-0.87) | 0.64 (0.52-0.79) | 0.61 (0.49-0.75) | <0.001 |
| Model 2 | 1.00<sup>a</sup> | 0.78 (0.64-0.96) | 0.73 (0.58-0.91) | 0.67 (0.53-0.84) | <0.001 |
| **uPDI** | **Q1** | **Q2** | **Q3** | **Q4** | **P-trend** |
| Cases/total | 151/1,250 | 177/1,374 | 157/1,148 | 190/1,035 |  |
| HR (95% CI) |  |  |  |  |  |
| Model 1 | 1.00<sup>a</sup> | 1.11 (0.89-1.38) | 1.20 (0.96-1.50) | 1.66 (1.34-2.07) | <0.001 |
| Model 2 | 1.00<sup>a</sup> | 1.07 (0.86-1.34) | 1.22 (0.96-1.54) | 1.49 (1.18-1.89) | 0.004 |

Model 1 adjusted for sex and education; stratified by age (5-year categories) and region.
Model 2: Model 1 plus BMI, waist circumference, ethnicity, physical activity, smoking status, alcohol intake, energy intake, multimorbidity index, polypharmacy, Townsend deprivation index, prevalent diabetes, total protein intake and number of completed dietary assessments.
P-trend is for linear trend.
<sup>a</sup>Reference categories.
Abbreviations: Q, quartile; hPDI, healthful plant-based diet index; uPDI, unhealthful plant-based diet index; BMI, Body Mass Index; HR, hazard ratio; CI, confidence interval.

Of the individual PDI components, higher intakes (Q4) of wholegrains (>2.9 servings/day) was associated with a 29% lower risk of mortality [HR_Q4vsQ1_ (95% CI): 0.71 (0.56-0.90), p_trend_ = 0.02] compared to lower intakes (Q1) (zero servings/day), while higher intakes (Q4) of refined grains (2.0 servings/day) and SSBs (1.1 serving/day) was associated with a 28% and 31% higher risk of mortality [1.28 (1.04-1.59), p_trend_ = 0.002; 1.31 (1.06-1.62), p_trend_ = 0.001], respectively, compared to lower intakes (Q1) (zero servings/day) (**Table 3**).

**Table 3.** Hazard ratios (95% confidence intervals) of mortality across key food groups from the plant-based diet index (n=4,807)

|  | Food group intake quartiles |  |  |  |  |
| --- | --- | --- | --- | --- | --- |
|  | Q1 | Q2 | Q3 | Q4 | P-trend |
| **Healthy Plant Foods** |  |  |  |  |  |
| **Wholegrains** |  |  |  |  |  |
| Intake (servings/d)<sup>a</sup> | 0 (0-0) | 1.0 (0.1-1.4) | 2.0 (1.5-2.9) | 3.8 (2.9-11.0) |  |
| Cases/total | 130/685 | 189/1,375 | 178/1,369 | 178/1,378 |  |
| HR (95% CI)<sup>b</sup> | 1.00<sup>c</sup> | 0.76 (0.60-0.96) | 0.82 (0.65-1.05) | 0.71 (0.56-0.90) | 0.02 |
| **Fruit** |  |  |  |  |  |
| Intake (servings/d)<sup>a</sup> | 0 (0-0) | 1.0 (0.1-1.4) | 2.0 (1.5-2.9) | 3.8 (2.9-18.0) |  |
| Cases/total | 105/582 | 164/1,161 | 213/1,610 | 193/1,454 |  |
| HR (95% CI)<sup>b</sup> | 1.00<sup>c</sup> | 0.88 (0.68-1.14) | 0.87 (0.68-1.12) | 0.86 (0.66-1.11) | 0.49 |
| **Vegetables** |  |  |  |  |  |
| Intake (servings/d)<sup>a</sup> | 0 (0-0) | 1.0 (0.1-1.6) | 2.3 (1.6-3.1) | 4.5 (3.1-24.0) |  |
| Cases/total | 108/667 | 203/1,385 | 192/1,429 | 172/1,326 |  |
| HR (95% CI)<sup>b</sup> | 1.00<sup>c</sup> | 1.08 (0.84-1.38) | 1.06 (0.82-1.36) | 1.05 (0.81-1.37) | 0.44 |
| **Nuts** |  |  |  |  |  |
| Intake (servings/d)<sup>a</sup> | 0 (0-0) | 0.3 (0.1-0.3) | 0.5 (0.4-0.7) | 1.0 (0.8-4.0) |  |
| Cases/total | 577/3,888 | 39/350 | 27/293 | 32/276 |  |
| HR (95% CI)<sup>b</sup> | 1.00<sup>c</sup> | 0.96 (0.68-1.35) | 0.69 (0.46-1.02) | 0.95 (0.66-1.37) | 0.44 |
| **Legumes** |  |  |  |  |  |
| Intake (servings/d)<sup>a</sup> | 0 (0-0) | 0.3 (0.1-0.4) | 0.5 (0.4-0.8) | 1.0 (0.8-6.0) |  |
| Cases/total | 326/2,227 | 102/795 | 103/855 | 144/930 |  |
| HR (95% CI)<sup>b</sup> | 1.00<sup>c</sup> | 0.96 (0.75-1.24) | 0.91 (0.71-1.15) | 0.99 (0.81-1.21) | 0.57 |
| **Tea and coffee** |  |  |  |  |  |
| Intake (servings/d)<sup>a</sup> | 0 (0-0) | 3.0 (0.3-3.8) | 4.5 (3.8-5.1) | 6.0 (5.1-18.0) |  |
| Cases/total | 13/108 | 245/1,573 | 230/1,745 | 187/1,381 |  |
| HR (95% CI)<sup>b</sup> | 1.00<sup>c</sup> | 1.16 (0.65-2.05) | 0.98 (0.55-1.75) | 0.97 (0.54-1.74) | 0.05 |
| **Unhealthy Plant Foods** |  |  |  |  |  |
| **Refined grains** |  |  |  |  |  |
| Intake (servings/d)<sup>a</sup> | 0 (0-0) | 0.5 (0.1-0.9) | 1.0 (1.0-1.9) | 2.8 (2.0-9.0) |  |
| Cases/total | 172/1,257 | 119/1,032 | 166/1,298 | 218/1,220 |  |
| HR (95% CI)<sup>b</sup> | 1.00<sup>c</sup> | 1.08 (0.83-1.40) | 0.97 (0.77-1.21) | 1.28 (1.04-1.59) | 0.002 |
| **Potatoes** |  |  |  |  |  |
| Intake (servings/d)<sup>a</sup> | 0 (0-0) | 0.5 (0.1-0.1) | 1.0 (0.6-1.0) | 1.5 (1.1-5.0) |  |
| Cases/total | 137/1,033 | 141/1,117 | 267/1,827 | 130/830 |  |
| HR (95% CI)<sup>b</sup> | 1.00<sup>c</sup> | 1.12 (0.87-1.44) | 1.13 (0.91-1.41) | 1.21 (0.93-1.57) | 0.38 |
| **Sugar-sweetened beverages** |  |  |  |  |  |
| Intake (servings/d)<sup>a</sup> | 0 (0-0) | 0.4 (0.1-0.6) | 1.0 (0.6-1.0) | 2.0 (1.1-7.0) |  |
| Cases/total | 318/2,524 | 97/759 | 118/758 | 142/766 |  |
| HR (95% CI)<sup>b</sup> | 1.00<sup>c</sup> | 1.09 (0.86-1.39) | 1.20 (0.97-1.50) | 1.31 (1.06-1.62) | 0.001 |
| **Fruit juices** |  |  |  |  |  |
| Intake (servings/d)<sup>a</sup> | 0 (0-0) | 0.3 (0.1-0.4) | 0.5 (0.5-0.9) | 1.0 (1.0-13.0) |  |
| Cases/total | 339/2,378 | 58/426 | 128/943 | 150/1,060 |  |
| HR (95% CI)<sup>b</sup> | 1.00<sup>c</sup> | 1.12 (0.83-1.52) | 1.09 (0.88-1.34) | 1.02 (0.84-1.25) | 0.55 |
| **Sweets and desserts** |  |  |  |  |  |
| Intake (servings/d)<sup>a</sup> | 0 (0-0) | 0.8 (0.1-1.1) | 1.7 (1.1-2.1) | 3.0 (2.1-9.0) |  |
| Cases/total | 131/930 | 177/1,525 | 172/1,177 | 195/1,175 |  |
| HR (95% CI)<sup>b</sup> | 1.00<sup>c</sup> | 0.88 (0.70-1.12) | 1.08 (0.84-1.38) | 1.10 (0.85-1.43) | 0.22 |
| **Animal-Derived Foods** |  |  |  |  |  |
| **Animal fat** |  |  |  |  |  |
| Intake (servings/d)<sup>a</sup> | 0 (0-0) | 0.8 (0.1-1.0) | 2.0 (1.1-2.0) | 3.0 (2.1-11.0) |  |
| Cases/total | 368/2,818 | 107/780 | 105/694 | 95/515 |  |
| HR (95% CI)<sup>b</sup> | 1.00<sup>c</sup> | 1.21 (0.97-1.52) | 1.04 (0.83-1.30) | 1.16 (0.92-1.48) | 0.14 |
| **Dairy** |  |  |  |  |  |
| Intake (servings/d)<sup>a</sup> | 0 (0-0) | 0.5 (0.1-0.9) | 1.0 (1.0-1.4) | 2.0 (1.5-7.0) |  |
| Cases/total | 133/834 | 150/1,246 | 195/1,301 | 197/1,426 |  |
| HR (95% CI)<sup>b</sup> | 1.00<sup>c</sup> | 0.88 (0.68-1.14) | 1.04 (0.83-1.32) | 0.91 (0.72-1.16) | 0.61 |
| **Eggs** |  |  |  |  |  |
| Intake (servings/d)<sup>a</sup> | 0 (0-0) | 0.3 (0.1-0.4) | 0.5 (0.5-0.9) | 1.0 (1.0-5.0) |  |
| Cases/total | 418/3,037 | 44/445 | 90/564 | 123/761 |  |
| HR (95% CI)<sup>b</sup> | 1.00<sup>c</sup> | 0.89 (0.63-1.25) | 1.24 (0.96-1.59) | 0.98 (0.80-1.22) | 0.39 |
| **Fish and seafood** |  |  |  |  |  |
| Intake (servings/d)<sup>a</sup> | 0 (0-0) | 0.3 (0.1-0.4) | 0.5 (0.4-0.8) | 1.0 (0.8-6.0) |  |
| Cases/total | 414/2,734 | 69/540 | 87/766 | 105/767 |  |
| HR (95% CI)<sup>b</sup> | 1.00<sup>c</sup> | 1.11 (0.82-1.49) | 0.91 (0.71-1.17) | 1.03 (0.82-1.29) | 0.94 |
| **Meat** |  |  |  |  |  |
| Intake (servings/d)<sup>a</sup> | 0 (0-0) | 0.5 (0.1-0.9) | 1.0 (1.0-1.4) | 2.0 (1.5-10.0) |  |
| Cases/total | 102/723 | 121/891 | 212/1,569 | 240/1,624 |  |
| HR (95% CI)<sup>b</sup> | 1.00<sup>c</sup> | 1.31 (0.98-1.75) | 1.17 (0.90-1.50) | 1.23 (0.94-1.61) | 0.14 |
| **Miscellaneous animal-based foods** |  |  |  |  |  |
| Intake (servings/d)<sup>a</sup> | 0 (0-0) | 0.3 (0.1-0.4) | 0.5 (0.5-0.8) | 1.0 (1.0-4.0) |  |
| Cases/total | 621/4,428 | 14/110 | 18/109 | 22/160 |  |
| HR (95% CI)<sup>b</sup> | 1.00<sup>c</sup> | 1.16 (0.67-2.02) | 1.45 (0.90-2.35) | 0.96 (0.62-1.49) | 0.63 |

<sup>a</sup>Intake values are median (range) and portion sizes were specified as a “serving” in the Oxford WebQ tool.
<sup>b</sup>Hazard Ratios with 95% Confidence Intervals (CI) adjusted for sex, BMI, waist circumference, ethnicity, physical activity, smoking status, education, alcohol intake, energy intake, multimorbidity index, polypharmacy, Townsend deprivation index, prevalent diabetes, total protein intake and number of completed dietary assessments; stratified by age (5-year categories) and region.
<sup>c</sup>Reference categories.
P-trend is for linear trend.
Abbreviations: Q, quartile; BMI, Body Mass Index; HR, hazard ratio; CI, confidence interval.

### 3.3 Subgroup and sensitivity analyses

For associations between hPDI and mortality, subgroup analysis showed no evidence of heterogeneity across key covariates (age, sex, smoking status, alcohol intake, protein intake, eGFR, PRS (eGFR), and CKD stage) **(Supplementary Table S8-S9)**. Results remained robust in sensitivity analyses: further adjusting for eGFR, genetic susceptibility of kidney diseases and excluding participants with short follow-up duration (<2 years) (**Supplementary Tables S10-S12**). Stronger associations were shown between hPDI and uPDI and mortality risk in repeat analyses restricting to ≥2 dietary assessments, although P_trend_ for uPDI turned non-significant (p_trend_ = 0.08) (**Supplementary Table S13**).

## 4. Discussion

In this large-scale cohort study, higher adherence to hPDI was associated with a 33% lower risk of all-cause mortality among individuals with CKD, while a higher uPDI was associated with a 49% higher risk following full adjustment for a range of diet and lifestyle factors. Associations between PDIs and mortality were only shown for CKD patients in early stages (stages 1-2).

Our findings support the role of adopting a healthy plant-based dietary pattern for lengthening survival of CKD. While it’s widely recognised that increased adherence to a PBD may lower mortality risk in the general population (31–33), to our knowledge, only one study currently exists on PBDs and risk of CKD progression and all-cause mortality in CKD patients from the Chronic Renal Insufficiency Cohort (CRIC) Study (34). While it’s reassuring to find that our results align with those from the CRIC study, one key difference to note is that our study consisted mostly of individuals with early stage CKD (97% with CKD stages 1-3b; mean (SD) eGFR of 59.5 (18.0)), while the CIRC study was predominantly made up of CKD patients with stages 3 and 4 (70% and 18% respectively; mean (SD) eGFR of 43.4 (13.5)) (35)). Collectively, these findings suggest that a healthful PBD exerts benefits across all stages of CKD, in line with previous studies to suggest that higher dietary intake of healthful plant foods lowers CKD risk through improved kidney function (36–38).

Several plausible biological mechanisms may explain the beneficial role of plant-rich diets on kidney health. Firstly, plant-rich diets are typically nutrient dense, providing a rich source of vitamins, minerals, and phytochemicals. Flavonoids, a group of secondary plant metabolites have shown to have renoprotective effects in CKD through their anti-inflammatory and antioxidant properties (39, 40). Supporting this, a randomized, double-blinded, placebo-controlled trial previously reported dietary cocoa flavanols to improve vascular function in patients with ESKD (41). Our study also revealed that individuals with a higher hPDI consumed 39% more dietary fibre than those with a low hPDI. A recent meta-analysis of 10 cohort studies indicated the potential protective effects of increasing dietary fibre consumption against all-cause mortality, cardiovascular mortality and cardiovascular disease in CKD patients (42). Moreover, previous research has demonstrated that increased dietary fibre intake is likely to reduce levels of inflammation in CKD patients (43–45). Results from an intervention trial demonstrated significantly lower levels of C-reactive protein, interleukin-6, interleukin-8, and tumour necrosis factor-α in haemodialysis patients supplemented with 10g and 20g of dietary fibre per day over 6 weeks (46). Further, through increased intestinal motility, dietary fibre has also been shown to play a favourable role in the gut microbiome, promoting the growth of beneficial microbiota, and decreasing uremic toxin production (47, 48). Supporting this, a meta-analysis of 14 controlled feeding trials revealed that greater dietary fibre intake was associated with lower serum urea and creatinine levels in individuals with CKD (49).

PBDs typically confer lower intakes of animal-based foods. Though the exact mechanisms through which animal foods such as red meat may contribute to renal toxicity are unclear, one proposed explanation is that increased consumption of animal foods leads to a high acid load in the diet (50). Higher acid load in the diet has been linked to renal toxicity and kidney function decline through metabolic acidosis in patients with CKD (51). It is also inferred that animal foods increase inflammation and promote unfavourable changes to the gut microbiome which lead to increased production of uremic toxins (52–54), potentially influencing the progression of renal disease. Although in this study, no significant associations were shown for higher meat intake and reduced mortality, evidence from the Nurses’ Health Study suggests that consuming two or more servings of red meat per week increases the risk of microalbuminuria (55), while the Singapore Chinese Health Study found a dose-dependent relationship between red meat intake and ESKD (56). In addition, a randomized crossover trial of postmenopausal women demonstrated that replacing meat with soy improved insulin sensitivity, offering a potential approach for managing and preventing metabolic syndrome (57)–a pathological state associated with decreased renal function (58, 59).

Our study also showed that higher intakes of SSBs were associated with increased mortality risk. While our findings align with those of a recent dose-response meta-analysis where greater consumption of SSBs was associated with a higher risk of all-cause mortality (60), numerous other studies have also observed that a higher intake of sugar or artificially sweetened beverages was associated with kidney function decline (61–63). Some possible biological mechanisms that may explain these associations include that SSBs contain high amounts of added sugar and lead to low satiety, contributing to weight gain and obesity (64, 65). Moreover, SSBs often contain high levels of fructose which has previously been linked to renal disease and its progression through hyperuricemia (66, 67) and kidney stone formation (68).

### 4.1 Limitations

There are some limitations of this study. Firstly, despite our efforts to account for various potential confounding factors, unmeasured or residual confounding could still be present given the observational design of the study, so results should be interpreted with caution. Secondly, the PDI was based on self-reported 24-hr dietary data, introducing the possibility of measurement error and recall bias. Thirdly, this study was confined to individuals with pre-existing CKD; to maximise the sample size, participants with data from at least one 24-h dietary assessment were included (2,118 (44.1%) of 4,807 completed one dietary assessment). While a single dietary assessment limits the reproducibility of food group level intakes, this study, in addition to others, has shown that diet quality scores including the PDIs are reproducible over time based on single dietary recalls (21, 69, 70). Fourthly, primary care data was not used to identify CKD cases, potentially constraining the sample size of this study. Despite this, eGFR was also used to classify patients with CKD, which is considered an optimal marker of kidney function (71). Finally, subgroup analyses by ethnicity were not conducted due to the limited number of non-white participants in the study (344 (7.2%) of 4,807), further highlighting that the UK Biobank is not representative of the UK population, limiting the generalisability of our findings (72).

## 5. Conclusion

In conclusion, better adherence to a healthy PBD was associated with a lower risk of all-cause mortality independent of a wide range of factors, including eGFR and genetic susceptibility to kidney diseases, while higher adherence to an unhealthy PBD was associated with a higher risk. While existing recommendations for nutritional management of CKD primarily target individual dietary components i.e., protein and sodium (71, 73), our data support the shift towards healthy plant-based dietary patterns, specifically through increasing intakes of wholegrains and lowering intakes of refined grains and sugar-sweetened beverages, to better manage and treat CKD patients. Our findings underscore the importance of conducting large-scale clinical trials to further explore the impact of plant-based dietary patterns on clinical outcomes among individuals with CKD.

### Funding statement

This research was conducted using UK Biobank funded and sourced data (application 64426). The UK Biobank was established by the Wellcome Trust, the Medical Research Council, the UK Department of Health, and the Scottish Government. The UK Biobank has also received funding from the Welsh Assembly Government, the British Heart Foundation, and Diabetes United Kingdom, Northwest Regional Development Agency, Scottish Government. In addition, Alysha S. Thompson holds a PhD studentship of the Department for the Economy (DfE), Northern Ireland. This work was in part supported by the Co-Centre for Sustainable and Resilient Food Systems.

### Conflict of interest

None were reported.

### Authors’ contributions

Design and concept: AT, TK, AC; database development: AT, ATR, JKO, CH; analysed and interpreted data: AT, AC, TK; drafted manuscript: AT, TK, AC; provided critical review of the manuscript: all authors; guarantors of the work: AT and TK.

### Data sharing statement

UK Biobank data can be requested by all bona fide researchers for approved projects, including replication, through https://www.ukbiobank.ac.uk/. This research was conducted using UK Biobank funded and sourced data (application 64426).

### Ethics statement

All UK Biobank participants provided informed consent to participate and be followed through linkage to their health records. The UK Biobank study received ethical approval from the NHS North West Multicentre Research Ethics Committee (Ref. 11/NW/0382).

Disclaimer: "The views expressed in this publication are those of the authors and should not be interpreted as representing the official position of the European Food Safety Authority (EFSA). Therefore, the present article is published under the sole responsibility of the authors, and may not be considered as an EFSA scientific output. EFSA cannot be held accountable for any errors or inaccuracies that may appear."

## Notes

### Competing Interest Statement

The authors have declared no competing interest.

