## Supplementary file 1 for "Adherence to a healthful plant-based diet and risk of mortality among individuals with chronic kidney disease: A prospective cohort study"

**Supplementary file 1 – Adherence to a healthful plant-based diet and risk of mortality among individuals with chronic kidney disease: A prospective cohort study**

***Thompson et al.***

### **Methods S1:** Genotyping and quality control

Additional quality control (QC) procedures were conducted using PLINK v2.0 (<https://www.cog-genomics.org/plink/2.0/>) (1, 2). These measures encompassed several thresholds. 1) Analyses were confined to individuals with self-reported White British ancestry and concordant self-reported and genetic sex. 2) Only autosomal variants were considered, with exclusion criteria set at a genotype call rate ≤95%, an imputation rate ≤97%, a minor allele frequency ≤0.001, a minimum count of minor alleles of 5, individual missingness ≤95%, and Hardy-Weinberg equilibrium ≤1e-8. 3) Outliers for heterozygosity and missingness (PCA corrected), individuals with suspected sex aneuploidy, and duplicates were identified and removed based on predefined metrics provided by the UKB (3). The final genetic UKB sample comprised 408,110 individuals, including 220,618 females and 187,492 males.

Included participants (n= 502,236)

All participants with chronic kidney disease (n= 31,556)

Total available participants with chronic kidney disease and data on ≥1 plausible dietary assessment (n=4,808)

1. Follow-up ended before completing final diet recall (n=1)

Final sample of chronic kidney disease patients (n=4,807) Total mortality cases (n=675)

1. Missing dietary data (n=26,670) 2. Implausible energy intakes (n=66) 3. Missing covariate information (n=12)

### **Figure S1.** Flowchart of exclusions

### **Table S1.** Codes used to ascertain chronic kidney disease

| **Outcome** | **Self-reported codes (data-field ID: 20002)** | **ICD-9 codes (data-field ID: 41271)** | **ICD-10 codes (data-field ID: 41270)** | **OPCS-3 codes (data-field ID: 41273)** | **OPCS-4 codes (data-field ID: 41272)** |
| --- | --- | --- | --- | --- | --- |
| **Chronic Kidney Disease** | 1193, 1194, 1192, 1607 | 585.9 | N03, N06, N08, N11, N12, N13, N14, N15, N16, N18, N19, Z49, I12, I13 | 560.2, 561, 561.1, 562, 563.1, 563.2, 563.3, 564, 564.1, 565, 566, 566.1, 567.1, 567.2, 568, 569, 571.1 | L74.1, L74.2, L74.3, L74.4, L74.5, L74.6, L74.8, L74.9, M01.2, M01.3, M01.4, M01.5, M01.8, M01.9, M02.3, M08.4, M17.2, M17.4, M17.8, M17.9, X40.1, X40.2, X40.3, X40.4, X40.5, X40.6, X40.7, X40.8, X40.9, X41.1, X41.2, X41.8, X41.9, X42.1, X42.8, X42.9, and X43.1 |

### **Table S2.** Genetic instruments for eGFR (n=161 SNPs)

| SNP | Chr | Pos_(b37) | Locus | EA | OA | EAF | Effect | SE | p-value |
| --- | --- | --- | --- | --- | --- | --- | --- | --- | --- |
| rs74748843 | 1 | 10730910 | CASZ1 | T | C | 0.07 | -0.0048 | 8e-4 | 3.7e-9 |
| rs10159261 | 1 | 15912987 | AGMAT | T | G | 0.36 | -0.0034 | 3e-4 | 4.8e-25 |
| rs12061708 | 1 | 18809916 | KLHDC7A | A | G | 0.29 | -0.0026 | 3e-4 | 9.6e-14 |
| rs659437 | 1 | 46037394 | AKR1A1 | T | C | 0.22 | -0.0027 | 4e-4 | 3.3e-12 |
| rs11211257 | 1 | 46581933 | PIK3R3 | A | G | 0.82 | 0.0027 | 5e-4 | 2.3e-9 |
| rs688540 | 1 | 48002447 | FOXD2 | A | G | 0.87 | -0.003 | 5e-4 | 3e-8 |
| rs17413465 | 1 | 55718708 | MIR4422HG | A | C | 0.18 | 0.0025 | 4e-4 | 8.9e-9 |
| rs1757915 | 1 | 56615809 | LINC01755 | A | G | 0.33 | 0.0021 | 3e-4 | 2.9e-10 |
| rs7536433 | 1 | 78023173 | AK5 | T | C | 0.26 | 0.0021 | 4e-4 | 6.7e-9 |
| rs679843 | 1 | 78707493 | MGC27382 | T | C | 0.33 | 0.0021 | 3e-4 | 5.2e-10 |
| rs1887252 | 1 | 82957871 | LINC01362 | C | G | 0.62 | -0.0019 | 3e-4 | 2.9e-9 |
| rs11166440 | 1 | 100808363 | CDC14A | A | G | 0.6 | 0.002 | 3e-4 | 1.8e-10 |
| rs10857788 | 1 | 110012289 | SYPL2 | A | G | 0.7 | 0.003 | 4e-4 | 2e-16 |
| rs12736457 | 1 | 113258293 | PPM1J | C | G | 0.87 | 0.0054 | 5e-4 | 1e-25 |
| rs267738 | 1 | 150940625 | CERS2 | T | G | 0.8 | -0.0048 | 4e-4 | 1.20e-32 |
| rs3845534 | 1 | 163738950 | LOC100422212 | A | G | 0.53 | -0.0019 | 3e-4 | 1.2e-9 |
| rs4656220 | 1 | 170649277 | PRRX1 | T | C | 0.42 | 0.002 | 3e-4 | 3.3e-10 |
| rs3795503 | 1 | 180905694 | KIAA1614 | T | C | 0.36 | 0.002 | 3e-4 | 9.8e-10 |
| rs78329830 | 1 | 186769572 | PLA2G4A | A | G | 0.96 | -0.0054 | 9e-4 | 6.7e-9 |
| rs3850625 | 1 | 201016296 | CACNA1S | A | G | 0.12 | 0.0046 | 5e-4 | 1.1e-18 |
| rs75625374 | 1 | 208039431 | CD34 | C | G | 0.06 | 0.0045 | 7e-4 | 4.5e-10 |
| rs7535253 | 1 | 214744893 | PTPN14 | T | C | 0.27 | 0.0021 | 4e-4 | 1.3e-9 |
| rs61830291 | 1 | 221001142 | LINC01352 | A | C | 0.9 | -0.0036 | 6e-4 | 1.2e-9 |
| rs417237 | 1 | 228532195 | OBSCN | T | G | 0.57 | 0.0018 | 3e-4 | 7.5e-9 |
| rs2490391 | 1 | 243469669 | SDCCAG8 | A | C | 0.43 | -0.0024 | 3e-4 | 1.3e-14 |
| rs3791221 | 2 | 226933 | SH3YL1 | A | G | 0.67 | 0.0022 | 3e-4 | 1.2e-11 |
| rs2301343 | 2 | 40680149 | SLC8A1 | T | G | 0.76 | -0.0023 | 4e-4 | 4.1e-10 |
| rs10865189 | 2 | 43433257 | ZFP36L2 | C | G | 0.51 | 0.0024 | 3e-4 | 3.3e-14 |
| rs2971880 | 2 | 54885640 | SPTBN1 | A | T | 0.37 | -0.0024 | 3e-4 | 7.6e-15 |
| rs10197255 | 2 | 67874553 | LINC01812 | A | T | 0.4 | 0.0018 | 3e-4 | 1.2e-8 |
| rs11694902 | 2 | 121988884 | TFCP2L1 | A | G | 0.14 | 0.0041 | 5e-4 | 1.1e-16 |
| rs7425436 | 2 | 148759656 | ORC4 | A | G | 0.65 | 0.0024 | 3e-4 | 5.2e-13 |
| rs35472707 | 2 | 169995581 | LRP2 | T | C | 0.05 | -0.0073 | 8e-4 | 6.2e-19 |
| rs187355703 | 2 | 176993583 | HOXD8 | C | G | 0.97 | 0.01 | 0.0011 | 1e-18 |
| rs4666821 | 2 | 183077254 | PDE1A | T | G | 0.53 | 0.002 | 3e-4 | 2.5e-11 |
| rs60980181 | 2 | 188168567 | CALCRL | A | T | 0.17 | -0.0027 | 4e-4 | 1.8e-10 |
| rs1047891 | 2 | 211540507 | CPS1 | A | C | 0.29 | -0.0065 | 4e-4 | 1.2e-75 |
| rs1548945 | 2 | 217665788 | TNP1 | T | C | 0.44 | 0.0036 | 3e-4 | 8.4e-31 |
| rs1050816 | 2 | 220358198 | SPEG | T | C | 0.33 | 0.0026 | 3e-4 | 1.1e-15 |
| rs35669853 | 2 | 227287718 | MIR5702 | A | G | 0.18 | 0.0024 | 4e-4 | 9.2e-9 |
| rs795009 | 3 | 12208671 | SYN2 | T | G | 0.73 | 0.002 | 3e-4 | 7e-9 |
| rs6778731 | 3 | 13947504 | WNT7A | T | C | 0.59 | -0.0017 | 3e-4 | 3.3e-8 |
| rs6779998 | 3 | 30749965 | TGFBR2 | A | G | 0.52 | -0.0017 | 3e-4 | 1.6e-8 |
| rs7651407 | 3 | 48443816 | PLXNB1 | T | C | 0.44 | 0.0025 | 4e-4 | 2.4e-11 |
| rs4625 | 3 | 49572140 | DAG1 | A | G | 0.7 | -0.0023 | 4e-4 | 5.5e-11 |
| rs2581820 | 3 | 53020544 | SFMBT1 | A | G | 0.29 | 0.0021 | 3e-4 | 7.9e-10 |
| rs2289746 | 3 | 105455955 | CBLB | T | C | 0.41 | -0.0019 | 3e-4 | 2.5e-9 |
| rs9868185 | 3 | 121657593 | SLC15A2 | A | G | 0.5 | 0.0026 | 3e-4 | 5e-17 |
| rs10934754 | 3 | 125906237 | ALDH1L1-AS2 | T | C | 0.6 | 0.002 | 3e-4 | 1.3e-10 |
| rs35320690 | 3 | 135932494 | MSL2 | T | C | 0.73 | -0.0025 | 4e-4 | 3e-11 |
| rs9828976 | 3 | 136536835 | SLC35G2 | C | G | 0.76 | -0.0024 | 4e-4 | 1.9e-9 |
| rs1397764 | 3 | 141750810 | TFDP2 | A | G | 0.27 | 0.0043 | 3e-4 | 2.5e-37 |
| rs76272256 | 3 | 168888112 | MECOM | T | C | 0.24 | 0.0024 | 4e-4 | 4.9e-10 |
| rs56065557 | 3 | 185354216 | SENP2 | C | G | 0.32 | -0.0029 | 3e-4 | 4.3e-18 |
| rs11919484 | 3 | 186432839 | KNG1 | T | G | 0.32 | -0.0026 | 3e-4 | 5.8e-16 |
| rs75501914 | 4 | 3449781 | HGFAC | A | G | 0.09 | 0.0039 | 6e-4 | 9e-11 |
| rs3775932 | 4 | 10090930 | WDR1 | A | C | 0.51 | -0.0018 | 3e-4 | 2e-9 |
| rs16874073 | 4 | 23743962 | PPARGC1A | T | C | 0.95 | -0.0045 | 7e-4 | 6.6e-11 |
| rs12509595 | 4 | 81182554 | FGF5 | T | C | 0.7 | -0.0035 | 3e-4 | 6.4e-25 |
| rs223471 | 4 | 103698786 | LOC102723704 | C | G | 0.34 | 0.0028 | 3e-4 | 5.9e-19 |
| rs55929207 | 4 | 109703549 | ETNPPL | C | G | 0.48 | 0.0019 | 3e-4 | 3.4e-10 |
| rs71606723 | 4 | 115498457 | UGT8 | A | T | 0.77 | 0.0025 | 4e-4 | 3.4e-12 |
| rs1362800 | 5 | 39378115 | DAB2 | T | C | 0.38 | -0.0049 | 3e-4 | 5.8e-51 |
| rs495237 | 5 | 39950266 | LINC00603 | T | G | 0.25 | 0.0027 | 3e-4 | 2e-14 |
| rs11746506 | 5 | 44812566 | MRPS30 | T | C | 0.41 | 0.0017 | 3e-4 | 3e-8 |
| rs12520984 | 5 | 52787358 | FST | C | G | 0.32 | 0.0019 | 3e-4 | 5.3e-9 |
| rs79760705 | 5 | 53298716 | ARL15 | T | G | 0.11 | 0.0056 | 5e-4 | 6.5e-25 |
| rs72759880 | 5 | 67750213 | PIK3R1 | T | G | 0.11 | -0.0056 | 5e-4 | 1.1e-26 |
| rs3797537 | 5 | 78322650 | DMGDH | A | G | 0.73 | 0.0019 | 3e-4 | 2.9e-8 |
| rs12777 | 5 | 131671662 | SLC22A4 | C | G | 0.96 | 0.005 | 9e-4 | 1.1e-8 |
| rs12163971 | 5 | 132226669 | AFF4 | A | C | 0.16 | -0.0029 | 4e-4 | 1.7e-12 |
| rs11743174 | 5 | 148524820 | ABLIM3 | T | C | 0.67 | 0.0019 | 3e-4 | 1.3e-8 |
| rs3812036 | 5 | 176813404 | SLC34A1 | T | C | 0.26 | -0.0065 | 4e-4 | 2.4e-74 |
| rs144100226 | 6 | 34180297 | HMGA1 | T | C | 0.04 | 0.0059 | 0.001 | 6e-9 |
| rs77915916 | 6 | 43287722 | CRIP3 | A | T | 0.92 | 0.0046 | 6e-4 | 7.3e-14 |
| rs720989 | 6 | 44765535 | SUPT3H | T | G | 0.79 | 0.0021 | 4e-4 | 1.8e-8 |
| rs12212034 | 6 | 51492862 | PKHD1 | T | C | 0.37 | -0.0018 | 3e-4 | 1e-8 |
| rs6458868 | 6 | 52630153 | GSTA2 | T | C | 0.67 | -0.002 | 3e-4 | 1.2e-9 |
| rs3925003 | 6 | 55422618 | HMGCLL1 | T | C | 0.58 | -0.0018 | 3e-4 | 2e-9 |
| rs72912510 | 6 | 90118764 | RRAGD | A | G | 0.2 | -0.0024 | 4e-4 | 6.4e-9 |
| rs1857859 | 6 | 100894587 | SIM1 | A | G | 0.31 | 0.0019 | 3e-4 | 2.6e-8 |
| rs7740107 | 6 | 130374461 | L3MBTL3 | A | T | 0.74 | 0.0027 | 4e-4 | 8.9e-13 |
| rs9375818 | 6 | 131882078 | ARG1 | A | G | 0.25 | -0.0031 | 4e-4 | 6.1e-18 |
| rs9397738 | 6 | 154986664 | SCAF8 | A | G | 0.84 | 0.0027 | 4e-4 | 3.5e-10 |
| rs12207180 | 6 | 160633107 | SLC22A2 | A | T | 0.11 | -0.0085 | 5e-4 | 2.6e-63 |
| rs6968554 | 7 | 17287106 | AHR | A | G | 0.43 | -0.0019 | 3e-4 | 1.1e-9 |
| rs700753 | 7 | 46753684 | LOC730338 | C | G | 0.32 | 0.0031 | 3e-4 | 2.1e-20 |
| rs55773927 | 7 | 65337902 | VKORC1L1 | T | C | 0.41 | 0.0019 | 3e-4 | 1.2e-8 |
| rs801193 | 7 | 66030612 | GS1-124K5.11 | T | G | 0.58 | -0.002 | 3e-4 | 1.9e-9 |
| rs41301394 | 7 | 75612803 | POR | T | C | 0.32 | 0.0023 | 3e-4 | 3.7e-12 |
| rs6973656 | 7 | 77422583 | TMEM60 | A | G | 0.64 | 0.0035 | 3e-4 | 5.70e-28 |
| rs3757387 | 7 | 128576086 | IRF5 | T | C | 0.59 | 0.003 | 3e-4 | 7e-20 |
| rs62491533 | 7 | 129564134 | UBE2H | T | C | 0.81 | -0.0027 | 4e-4 | 1.1e-11 |
| rs1533059 | 8 | 8684953 | MFHAS1 | A | G | 0.51 | 0.0025 | 3e-4 | 1.2e-14 |
| rs10098664 | 8 | 11417493 | BLK | T | C | 0.49 | -0.0021 | 3e-4 | 6.1e-10 |
| rs2976178 | 8 | 87332552 | WWP1 | C | G | 0.67 | -0.0025 | 3e-4 | 7.8e-14 |
| rs2954017 | 8 | 126476873 | TRIB1 | T | C | 0.46 | 0.0024 | 3e-4 | 1.7e-12 |
| rs12377027 | 9 | 20554583 | MLLT3 | A | G | 0.82 | -0.0026 | 5e-4 | 2.9e-8 |
| rs13287724 | 9 | 33169034 | B4GALT1-AS1 | A | T | 0.89 | -0.003 | 6e-4 | 4.7e-8 |
| rs2039424 | 9 | 71432174 | PIP5K1B | A | G | 0.64 | 0.0044 | 3e-4 | 2.1e-44 |
| rs1321917 | 9 | 119324929 | ASTN2 | C | G | 0.44 | -0.0023 | 3e-4 | 1.4e-13 |
| rs7024579 | 9 | 139100413 | QSOX2 | T | C | 0.29 | 0.0023 | 4e-4 | 8.2e-11 |
| rs80282103 | 10 | 899071 | LARP4B | A | T | 0.91 | 0.0078 | 6e-4 | 1.2e-44 |
| rs6481598 | 10 | 29781798 | SVIL | C | G | 0.79 | 0.0024 | 4e-4 | 1.5e-9 |
| rs10821905 | 10 | 52646093 | A1CF | A | G | 0.18 | 0.0037 | 4e-4 | 9.4e-19 |
| rs10821944 | 10 | 63785089 | ARID5B | T | G | 0.7 | 0.002 | 3e-4 | 3.9e-9 |
| rs7475348 | 10 | 69965177 | MYPN | T | C | 0.46 | 0.0031 | 3e-4 | 1.2e-22 |
| rs816850 | 10 | 79252446 | KCNMA1 | C | G | 0.25 | -0.002 | 4e-4 | 7.4e-9 |
| rs7095954 | 10 | 82209232 | TSPAN14 | A | T | 0.45 | -0.0018 | 3e-4 | 3.7e-8 |
| rs4918943 | 10 | 97278922 | SORBS1 | A | G | 0.22 | -0.0022 | 4e-4 | 9.2e-9 |
| rs284859 | 10 | 104573017 | WBP1L | T | G | 0.21 | 0.0026 | 4e-4 | 5e-12 |
| rs1055256 | 10 | 126446592 | EEF1AKMT2 | A | G | 0.42 | 0.0025 | 3e-4 | 3.6e-16 |
| rs11564722 | 11 | 2178330 | INS-IGF2 | T | C | 0.31 | 0.0033 | 4e-4 | 2.1e-20 |
| rs63934 | 11 | 2789062 | KCNQ1 | A | G | 0.83 | 0.0041 | 4e-4 | 3.90e-23 |
| rs61897431 | 11 | 47427667 | SLC39A13 | T | C | 0.65 | 0.0029 | 4e-4 | 6.3e-16 |
| rs948493 | 11 | 65552154 | MIR1234 | T | C | 0.33 | -0.0033 | 3e-4 | 2e-24 |
| rs6589750 | 11 | 119326726 | USP2-AS1 | A | G | 0.63 | 0.002 | 3e-4 | 1.5e-9 |
| rs117113238 | 12 | 12209203 | BCL2L14 | A | G | 0.09 | 0.0039 | 6e-4 | 8.6e-11 |
| rs41284816 | 13 | 50655989 | DLEU2 | T | G | 0.03 | -0.0078 | 0.0012 | 1.7e-10 |
| rs72683923 | 14 | 50735947 | L2HGDH | T | C | 0.98 | -0.0074 | 0.0013 | 3.4e-8 |
| rs1028455 | 14 | 88829975 | SPATA7 | A | T | 0.33 | 0.002 | 3e-4 | 4.8e-10 |
| rs17184313 | 14 | 93102251 | RIN3 | T | C | 0.17 | -0.0029 | 5e-4 | 2e-10 |
| rs61993680 | 14 | 100752644 | SLC25A29 | A | C | 0.61 | -0.0019 | 3e-4 | 1.5e-8 |
| rs12913015 | 15 | 39305443 | C15orf54 | T | C | 0.41 | 0.0027 | 3e-4 | 2.3e-17 |
| rs1994887 | 15 | 57793765 | CGNL1 | A | C | 0.26 | -0.002 | 4e-4 | 1.6e-8 |
| rs956006 | 15 | 62808539 | MGC15885 | T | C | 0.33 | 0.0019 | 3e-4 | 4.4e-9 |
| rs11071939 | 15 | 67463391 | SMAD3 | T | C | 0.93 | -0.0039 | 6e-4 | 4.9e-10 |
| rs4886755 | 15 | 76298132 | NRG4 | A | G | 0.5 | 0.0041 | 3e-4 | 2e-39 |
| rs17507300 | 15 | 83722059 | BTBD1 | A | G | 0.83 | 0.0024 | 4e-4 | 1e-8 |
| rs7169629 | 15 | 85191274 | WDR73 | C | G | 0.48 | 0.0018 | 3e-4 | 1.6e-8 |
| rs59646751 | 15 | 99276521 | IGF1R | T | G | 0.3 | -0.0023 | 3e-4 | 3.1e-12 |
| rs438339 | 16 | 2003425 | RPL3L | T | C | 0.88 | 0.0035 | 6e-4 | 5e-8 |
| rs77924615 | 16 | 20392332 | PDILT | A | G | 0.2 | 0.0098 | 4e-4 | 1.5e-138 |
| rs9932625 | 16 | 51735746 | LINC01571 | A | G | 0.26 | -0.003 | 3e-4 | 2.2e-17 |
| rs7203398 | 16 | 53189672 | CHD9 | A | C | 0.74 | 0.0025 | 3e-4 | 4.7e-13 |
| rs62050038 | 16 | 69802865 | WWP2 | A | T | 0.83 | 0.0028 | 4e-4 | 1.3e-11 |
| rs62053077 | 16 | 71643669 | MARVELD3 | T | G | 0.43 | -0.0021 | 4e-4 | 3.7e-9 |
| rs1858800 | 16 | 73024276 | ZFHX3 | T | C | 0.32 | 0.002 | 3e-4 | 2.1e-9 |
| rs28581385 | 16 | 79942679 | LINC01229 | A | T | 0.84 | -0.0028 | 4e-4 | 1.4e-11 |
| rs2440165 | 17 | 19428719 | SLC47A1 | T | C | 0.64 | 0.004 | 3e-4 | 1.70e-31 |
| rs35662455 | 17 | 56755223 | TEX14 | C | G | 0.89 | 0.003 | 5e-4 | 3.9e-8 |
| rs883541 | 17 | 66449122 | PRKAR1A | A | G | 0.72 | -0.0022 | 3e-4 | 2.7e-10 |
| rs1719934 | 18 | 5585158 | EPB41L3 | A | G | 0.58 | 0.0026 | 3e-4 | 2.6e-17 |
| rs16942751 | 18 | 24393213 | AQP4 | A | C | 0.18 | -0.0029 | 5e-4 | 2.2e-9 |
| rs2974751 | 19 | 13053034 | CALR | A | C | 0.38 | 0.0018 | 3e-4 | 4.4e-8 |
| rs8101667 | 19 | 33402419 | CEP89 | T | C | 0.39 | 0.0044 | 3e-4 | 9.30e-44 |
| rs34647824 | 19 | 50138143 | RRAS | A | C | 0.74 | -0.0021 | 4e-4 | 4e-8 |
| rs62187537 | 20 | 1333060 | FKBP1A-SDCBP2 | T | C | 0.07 | 0.0039 | 7e-4 | 9.2e-9 |
| rs1041606 | 20 | 14677788 | MACROD2 | T | C | 0.23 | -0.0021 | 4e-4 | 2.5e-8 |
| rs6087579 | 20 | 32985155 | ITCH | A | G | 0.48 | -0.0028 | 3e-4 | 1.4e-19 |
| rs2235826 | 20 | 56143169 | PCK1 | A | T | 0.79 | -0.003 | 4e-4 | 6.8e-15 |
| rs1407040 | 20 | 57472174 | GNAS | T | C | 0.68 | 0.0018 | 3e-4 | 1.4e-8 |
| rs35636653 | 20 | 60858758 | OSBPL2 | T | C | 0.34 | 0.0022 | 3e-4 | 2.3e-11 |
| rs4408777 | 20 | 62706105 | RGS19 | A | G | 0.51 | -0.0021 | 3e-4 | 5.4e-11 |
| rs2823139 | 21 | 16576783 | NRIP1 | A | G | 0.33 | -0.0026 | 3e-4 | 5.2e-16 |
| rs2834317 | 21 | 35356706 | LOC101928126 | A | G | 0.14 | -0.0035 | 5e-4 | 4.3e-14 |
| rs2244237 | 21 | 37818141 | CLDN14 | T | G | 0.22 | 0.0027 | 4e-4 | 6.3e-11 |
| rs80576 | 22 | 36539804 | APOL3 | A | G | 0.16 | -0.0028 | 5e-4 | 1.3e-9 |
| rs4820324 | 22 | 38599857 | MAFF | C | G | 0.59 | -0.0023 | 3e-4 | 5.1e-14 |
| rs112880707 | 22 | 40884662 | MKL1 | T | C | 0.17 | 0.0052 | 5e-4 | 4.90e-31 |
| rs738527 | 22 | 43112961 | A4GALT | T | C | 0.29 | 0.0032 | 3e-4 | 4.2e-21 |
| *Abbreviations: SNP, single nucleotide polymorphism; Chr, chromosome; Pos_(b37), position and human genome reference build; EA, effect allele; OA, other allele; EAF, effect allele frequency; SE, standard error.* | | | | | | | | | |

| **Table S3.** Baseline characteristics across quartiles (Q) of the unhealthful plant-based diet index in the UK Biobank (n=4,807) | | | | | | |
| --- | --- | --- | --- | --- | --- | --- |
|  | **Participants, No. (%)^a^** | | | | | |
| **Characteristics across uPDI** | **Q1** | **Q2** | **Q3** | | **Q4** | **Whole sample** |
| **Number of participants** | 1,250 (26.0) | 1,374 (28.6) | 1,148 (23.9) | | 1,035 (21.5) | 4,807 (100.0) |
| **Mortality cases** | 151 (12.1) | 177 (12.9) | 157 (13.7) | | 190 (18.4) | 675 (14.0) |
| **Unhealthful plant-based diet index, mean (SD)** | 47.9 (1.5) | 53.8 (1.5) | 58.0 (1.2) | | 63.4 (2.9) | 55.3 (6.0) |
| **Sex-Female** | 615 (49.2) | 761 (55.4) | 567 (49.4) | | 509 (49.2) | 2,452 (51.0) |
| **Age at recruitment (years), mean (SD)** | 61.6 (6.4) | 61.2 (6.7) | 60.7 (6.9) | | 60.1 (7.5) | 60.9 (6.9) |
| **BMI (kg/m^2^), mean (SD)** | 28.3 (4.9) | 28.4 (5.3) | 28.5 (5.0) | | 29.1 (5.5) | 28.5 (5.2) |
| **Waist circumference (cm), mean (SD)** | 93.7 (13.9) | 93.2 (14.4) | 93.8 (13.7) | | 95.8 (14.3) | 94.0 (14.1) |
| **Energy intake (kJ/day), mean (SD)** | 8876.6 (2169.5) | 8392.4 (2135.9) | | 8063.2 (2229.5) | 8008.7 (2304.2) | 8357.1 (2229.9) |
| **Physical activity (MET-h/wk), mean (SD)** | 30.9 (39.0) | 30.3 (39.0) | 29.4 (41.8) | | 28.6 (45.8) | 29.9 (41.2) |
| **Ethnicity** |  | | | | |  |
| Asian | 51 (4.1) | 46 (3.4) | 43 (3.8) | | 33 (3.2) | 173 (3.6) |
| Black | 6 (0.5) | 0 (0.0) | 5 (0.4) | | 2 (0.2) | 13 (0.3) |
| Multiple | 35 (2.8) | 39 (2.8) | 44 (3.8) | | 40 (3.9) | 158 (3.3) |
| White | 1,145 (91.6) | 1,268 (92.3) | 1,040 (90.6) | | 949 (91.7) | 4,402 (91.6) |
| Other/missing^b^ | 13 (1.0) | 21 (1.5) | 16 (1.4) | | 11 (1.1) | 61 (1.3) |
| **Education^c^** |  |  |  | |  |  |
| Low | 288 (23.0) | 322 (23.4) | 271 (23.6) | | 271 (26.2) | 1,152 (24.0) |
| Medium | 198 (15.8) | 228 (16.6) | 192 (16.7) | | 169 (16.3) | 787 (16.4) |
| High | 629 (50.3) | 622 (45.3) | 480 (41.8) | | 385 (37.2) | 2,116 (44.0) |
| Missing | 135 (10.8) | 202 (14.7) | 205 (17.9) | | 210 (20.3) | 752 (15.6) |
| **Smoking status** |  | | | | |  |
| Never | 621 (49.7) | 737 (53.6) | 605 (52.7) | | 541 (52.3) | 2,504 (52.1) |
| Previous | 571 (45.7) | 558 (40.6) | 460 (40.1) | | 401 (38.7) | 1,990 (41.4) |
| Current | 53 (4.2) | 77 (5.6) | 78 (6.8) | | 91 (8.8) | 299 (6.2) |
| Missing | 5 (0.4) | 2 (0.2) | 5 (0.4) | | 2 (0.2) | 14 (0.3) |
| **Alcohol intake (g/day), mean (SD)** | 14.4 (19.2) | 13.1 (18.6) | 13.3 (19.8) | | 11.8 (19.7) | 13.2 (19.3) |
| **Prevalent diabetes** | 137 (11.0) | 155 (11.3) | 127 (11.1) | | 124 (12.0) | 543 (11.3) |
| **Multimorbidity** |  |  |  | |  |  |
| 0 LTCs | 358 (28.6) | 395 (28.8) | 343 (30.0) | | 275 (26.6) | 1,371 (28.5) |
| 1 LTC | 318 (25.4) | 359 (26.1) | 306 (26.7) | | 259 (25.0) | 1,242 (25.8) |
| 2 LTCs | 308 (24.6) | 319 (23.2) | 259 (22.6) | | 234 (22.6) | 1,120 (23.3) |
| ≥3 LTCs | 266 (21.3) | 301 (21.9) | 240 (20.9) | | 267 (25.8) | 1,074 (22.3) |
| **eGFR (mL/min/1.73 m2), mean (SD)** | 60.4 (17.2) | 59.3 (17.7) | 59.1 (17.9) | | 59.1 (19.4) | 59.5 (18.0) |
| **Protein intake (g/day), mean (SD)** | 90.4 (24.1) | 81.1 (21.3) | 74.7 (21.7) | | 70.5 (22.6) | 79.7 (23.6) |
| **uPDI food item intake (portion/day)^d^, mean (SD)** |  |  |  | |  |  |
| **Healthy plant food** |  |  |  | |  |  |
| Whole grains | 2.9 (1.7) | 2.2 (1.6) | 1.7 (1.4) | | 1.1 (1.3) | 2.0 (1.6) |
| Fruit | 2.9 (1.7) | 2.4 (1.7) | 1.9 (1.5) | | 1.3 (1.3) | 2.2 (1.7) |
| Vegetables | 3.4 (2.4) | 2.5 (2.1) | 1.9 (1.8) | | 1.3 (1.5) | 2.4 (2.2) |
| Nuts | 0.2 (0.5) | 0.1 (0.4) | 0.1 (0.3) | | 0.4 (0.2) | 0.1 (0.4) |
| Legumes | 0.6 (0.6) | 0.4 (0.5) | 0.3 (0.5) | | 0.2 (0.5) | 0.4 (0.5) |
| Tea and coffee | 4.9 (1.7) | 4.4 (1.8) | 4.1 (1.8) | | 3.6 (1.8) | 4.3 (1.8) |
| **Unhealthy plant food** |  |  |  | |  |  |
| Refined grains | 0.7 (1.0) | 1.1 (1.2) | 1.3 (1.3) | | 1.8 (1.5) | 1.2 (1.3) |
| Potatoes | 0.6 (0.5) | 0.7 (0.6) | 0.8 (0.7) | | 0.9 (0.7) | 0.7 (0.6) |
| Sugar-sweetened beverages | 0.3 (0.7) | 0.5 (0.8) | 0.6 (0.9) | | 1.1 (1.2) | 0.6 (1.0) |
| Fruit juices | 0.3 (0.5) | 0.4 (0.6) | 0.5 (0.7) | | 0.5 (0.7) | 0.4 (0.6) |
| Sweets and desserts | 1.1 (1.1) | 1.4 (1.3) | 1.6 (1.4) | | 2.0 (1.6) | 1.5 (1.4) |
| **Animal-derived food** |  |  |  | |  |  |
| Animal fat | 1.0 (1.4) | 0.8 (1.2) | 0.7 (1.2) | | 0.6 (1.0) | 0.8 (1.2) |
| Dairy | 1.3 (0.9) | 1.1 (0.8) | 0.9 (0.8) | | 0.8 (0.8) | 1.1 (0.9) |
| Eggs | 0.5 (0.6) | 0.3 (0.5) | 0.2 (0.5) | | 0.2 (0.4) | 0.3 (0.5) |
| Fish or seafood | 0.4 (0.5) | 0.3 (0.5) | 0.3 (0.4) | | 0.2 (0.4) | 0.3 (0.5) |
| Meat | 1.3 (1.0) | 1.3 (1.0) | 1.2 (1.0) | | 1.1 (1.0) | 1.2 (1.0) |
| Miscellaneous animal-based foods | 0.1 (0.3) | 0.1 (0.3) | 0.1 (0.3) | | 0.1 (0.3) | 0.1 (0.3) |
| ^a^Relative frequencies (%) include missing values which may not equate to 100%.  ^b^Other includes any race or ethnic group not otherwise specified.  ^c^Education was categorised as Low: CSEs or equivalent, O levels/GCSEs or equivalent; Medium: A levels/AS levels or equivalent, NVQ or HND or HNC or equivalent; High: College or University degree, other professional qualifications eg: nursing, teaching.  ^d^Portion sizes were specified as a “serving” in the Oxford WebQ tool.  *Abbreviations: Q, quartile; uPDI, unhealthful plant-based diet index; BMI, body mass index; MET, metabolic equivalent task; SD, standard deviation; LTC, long term condition; eGFR, estimated glomerular filtration rate.* | | | | | | |

| **Key nutrient intakes** |  | **Mean (SD)** | |  |
| --- | --- | --- | --- | --- |
| **Healthful plant-based diet index quartiles** | **Q1** | **Q2** | **Q3** | **Q4** |
| Participants, No. (%) | 1,309 (27.2) | 1,239 (25.8) | 1,131 (25.5) | 1,128 (23.5) |
| Healthful plant-based diet index | 49.7 (3.2) | 55.6 (1.5) | 59.5 (1.5) | 64.9 (2.7) |
| Energy, kJ/day | 9125.7 (2298.2) | 8370.9 (2107.8) | 7997.0 (2139.7) | 7810.8 (2118.5) |
| Protein, g/day | 83.9 (25.0) | 79.6 (22.6) | 77.7 (23.6) | 76.9 (22.4) |
| Fibre, g/day | 14.8 (5.4) | 16.3 (5.7) | 17.6 (6.0) | 20.6 (6.9) |
| Saturated fat, g/day | 32.1 (11.7) | 27.4 (10.7) | 24.4 (9.8) | 21.5 (9.6) |
| Cholesterol, mg/day | 310.2 (191.3) | 261.5 (158.6) | 228.2 (145.1) | 196.6 (129.1) |
| Glucose, g/day | 23.4 (11.8) | 24.5 (12.6) | 25.6 (13.1) | 28.3 (13.7) |
| Sodium, mg/day | 2253.8 (807.4) | 1949.0 (694.6) | 1779.2 (654.8) | 1661.6 (667.4) |
| Potassium, mg/day | 3385.2 (993.2) | 3485.5 (1001.1) | 3590.7 (1042.4) | 3810.3 (1043.2) |
| *Abbreviations: SD, standard deviation; Q, quartile.* | | | | |

### **Table S4.** Key nutrient intakes across quartiles (Q) of healthful plant-based diet index (N=4,807)

| **Key nutrient intakes** |  | **Mean (SD)** | |  |
| --- | --- | --- | --- | --- |
| **Unhealthful plant-based diet index quartiles** | **Q1** | **Q2** | **Q3** | **Q4** |
| Participants, No. (%) | 1,250 (26.0) | 1,374 (28.6) | 1,148 (23.9) | 1,035 (21.5) |
| Unhealthful plant-based diet index | 47.9 (1.5) | 53.8 (1.5) | 58.0 (1.2) | 63.4 (2.9) |
| Energy, kJ/day | 8876.6 (2169.5) | 8392.4 (2135.9) | 8063.2 (2229.5) | 8008.7 (2304.2) |
| Protein, g/day | 90.4 (24.1) | 81.1 (21.3) | 74.7 (21.7) | 70.5 (22.6) |
| Fibre, g/day | 20.8 (6.5) | 17.9 (5.8) | 15.7 (5.2) | 13.6 (5.6) |
| Saturated fat, g/day | 28.4 (11.6) | 26.8 (10.8) | 25.5 (11.3) | 25.4 (11.1) |
| Cholesterol, mg/day | 306.4 (184.7) | 258.4 (164.4) | 224.4 (147.6) | 201.6 (135.1) |
| Glucose, g/day | 28.9 (13.2) | 26.1 (13.0) | 23.7 (11.8) | 21.9 (12.3) |
| Sodium, mg/day | 2122.8 (765.8) | 1949.2 (732.4) | 1819.3 (722.2) | 1769.5 (713.3) |
| Potassium, mg/day | 4050.5 (973.9) | 3652.7 (979.8) | 3361.7 (993.0) | 3060.7 (977.1) |
| *Abbreviations: SD, standard deviation; Q, quartile.* | | | | |

### **Table S5.** Key nutrient intakes across quartiles (Q) of unhealthful plant-based diet index (N=4,807)

|  | **Participants, No. (%)^a^** | |
| --- | --- | --- |
| **Characteristics** | **Dietary data (N=4,807)** | **No dietary data (N=26,670)** |
| **Mortality cases** | 675 (14.0) | 6,200 (23.3) |
| **Age at recruitment (years), mean (SD)** | 60.9 (6.9) | 61.5 (6.7) |
| **BMI (kg/m^2^), mean (SD)** | 28.5 (5.2) | 29.5 (5.4) |
| **Waist circumference (cm), mean (SD)** | 94.0 (14.1) | 96.4 (14.3) |
| **Education** |  |  |
| Low | 1,152 (24.0) | 5,428 (20.4) |
| Medium | 787 (16.4) | 3,880 (14.6) |
| High | 2,116 (44.0) | 7,001 (26.4) |
| Missing | 752 (15.6) | 10,361 (38.9) |
| **Smoking status** |  |  |
| Never | 2,504 (52.1) | 12,254 (46.0) |
| Previous | 1,990 (41.4) | 10,779 (40.4) |
| Current | 299 (6.2) | 3,340 (12.5) |
| Missing | 14 (0.3) | 297 (1.1) |
| **eGFR (mL/min/1.73 m2), mean (SD)** | 59.5 (18.0) | 74.0 (19.2) |
| **CKD Stage** |  |  |
| 1-2 | 1,145 (27.2) | 17,794 (71.9) |
| 3a-3b | 2,628 (62.3) | 5,719 (23.1) |
| 3-5 | 443 (10.5) | 1,247 (5.0) |
| ^a^Relative frequencies (%) include missing values which may not equate to 100%. *Abbreviations: SD, standard deviation; BMI, body mass index; eGFR, estimated glomerular filtration rate; CKD, chronic kidney disease* | | |

### **Table S6.** Comparison of baseline characteristics of chronic kidney disease patients with and without 24-hr dietary data

| **Table S7.** Intraclass coefficients for hPDI and uPDI over time | | | | |
| --- | --- | --- | --- | --- |
|  | **24-hr dietary assessment cycles** | | | |
|  | T0 *vs.* T1 | T1 *vs.* T2 | T2 *vs.* T3 | T3 *vs.* T4 |
| **Participants, No.** | 428 | 1,075 | 1,132 | 1,290 |
| hPDI | 0.49 | 0.48 | 0.49 | 0.53 |
| **Participants, No.** | 428 | 1,075 | 1,132 | 1,290 |
| uPDI | 0.42 | 0.45 | 0.43 | 0.48 |
| *Abbreviations: hPDI, healthful plant-based diet index; uPDI, unhealthful plant-based diet index; T0, timepoint 0 (initial assessment visit); T1, timepoint 1 (on-line cycle 1); T2, timepoint 2 (on-line cycle 2); T3, timepoint 3 (on-line cycle 3); T4, timepoint 4 (on-line cycle 4).* | | | | |

| **Table S8.** Healthful plant-based diet score and mortality risk stratified by UK Biobank population subgroups | | | | |
| --- | --- | --- | --- | --- |
|  | **Cases/ total** | **hPDI (10-point increments)** | **P-trend** | **P- interaction** |
| **Age, years** |  |  |  |  |
| HR (95% CI)^a^ |  |  |  | 0.19 |
| <60 | 205/2,352 | 0.68 (0.53-0.87) | 0.003 |  |
| ≥63 | 470/2,455 | 0.80 (0.68-0.94) | 0.008 |  |
| **Sex** |  |  |  |  |
| HR (95% CI)^a^ |  |  |  | 0.43 |
| Male | 432/2,355 | 0.78 (0.65-0.93) | 0.005 |  |
| Female | 243/2,452 | 0.74 (0.58-0.93) | 0.01 |  |
| **Smoking status** |  |  |  |  |
| HR (95% CI)^a^ |  |  |  | 0.85 |
| Never | 256/ 2,504 | 0.73 (0.58-0.92) | 0.008 |  |
| Ever | 417/2,289 | 0.76 (0.64-0.91) | 0.003 |  |
| **Alcohol intake, g/day** |  |  |  |  |
| HR (95% CI)^a^ |  |  |  | 0.60 |
| Low | 247/ 1,662 | 0.74 (0.59-0.93) | 0.009 |  |
| Moderate | 213/1,543 | 0.70 (0.54-0.90) | 0.006 |  |
| High | 215/1,602 | 0.88 (0.68-1.14) | 0.32 |  |
| **Total protein, g/day** |  |  |  |  |
| HR (95% CI)^a^ |  |  |  | 0.41 |
| <median | 347/2,404 | 0.68 (0.56-0.83) | <0.001 |  |
| ≥median | 328/2,403 | 0.81 (0.67-0.99) | 0.04 |  |
| **eGFR, mL/min/1.73 m2** |  |  |  |  |
| HR (95% CI)^a^ |  |  |  | 0.92 |
| <35 | 74/208 | 0.44 (0.21-0.93) | 0.03 |  |
| ≥35 | 601/4,599 | 0.76 (0.65-0.88) | <0.001 |  |
| **PRS (eGFR)** |  |  |  |  |
| HR (95% CI)^a^ |  |  |  | 0.53 |
| Low | 182/1,326 | 0.68 (0.51-0.91) | 0.01 |  |
| Medium | 195/1,326 | 0.69 (0.53-0.89) | 0.004 |  |
| High | 187/1,326 | 0.90 (0.69-1.18) | 0.44 |  |
| **CKD stage** |  |  |  |  |
| HR (95% CI)^a^ |  |  |  | 0.07 |
| 1-2 | 124/1,063 | 0.57 (0.41-0.79) | 0.001 |  |
| 3a-3b | 365/3,071 | 0.91 (0.75-1.10) | 0.32 |  |
| 4-5 | 144/515 | 0.97 (0.68-1.37) | 0.85 |  |
| ^a^Hazard Ratios with 95% Confidence Intervals (CI), adjusted for sex (excluding subgroup analysis), BMI, waist circumference, ethnicity, physical activity, smoking status (excluding subgroup analysis), education, alcohol intake (excluding subgroup analysis), energy intake, multimorbidity index, polypharmacy, Townsend deprivation index, prevalent diabetes, total protein intake (excluding subgroup analysis) and number of completed dietary assessments; stratified by age (5-year categories) (excluding subgroup analysis) and region.  Heterogeneity was tested by comparing two models – one without an interaction term between subgroup of interest and hPDI (categorical), with a model that included an interaction term. The likelihood ratio test was used to produce P-interaction values.  *Abbreviations: Q, quartile; hPDI, healthful plant-based diet index; BMI, Body Mass Index; HR, hazard ratio; CI, confidence interval; eGFR, estimated glomerular filtration rate; PRS, polygenic risk score; CKD, chronic kidney disease.* | | | | |

| **Table S9.** Unhealthful plant-based diet score and mortality risk stratified by UK Biobank population subgroups | | | | |
| --- | --- | --- | --- | --- |
|  | **Cases/ total** | **hPDI (10-point increments)** | **P-trend** | **P- interaction** |
| **Age, years** |  |  |  |  |
| HR (95% CI)^a^ |  |  |  | 0.84 |
| <60 | 205/2,352 | 1.13 (0.88-1.47) | 0.34 |  |
| ≥63 | 470/2,455 | 1.27 (1.07-1.50) | 0.006 |  |
| **Sex** |  |  |  |  |
| HR (95% CI)^a^ |  |  |  | 0.39 |
| Male | 432/2,355 | 1.12 (0.93-1.34) | 0.23 |  |
| Female | 243/2,452 | 1.44 (1.14-1.81) | 0.003 |  |
| **Smoking status** |  |  |  |  |
| HR (95% CI)^a^ |  |  |  | 0.75 |
| Never | 256/ 2,504 | 1.26 (1.00-1.60) | 0.06 |  |
| Ever | 417/2,289 | 1.23 (1.02-1.47) | 0.03 |  |
| **Alcohol intake, g/day** |  |  |  |  |
| HR (95% CI)^a^ |  |  |  | 0.85 |
| Low | 247/ 1,662 | 1.31 (1.05-1.64) | 0.02 |  |
| Moderate | 213/1,543 | 1.40 (1.07-1.82) | 0.01 |  |
| High | 215/1,602 | 1.02 (0.77-1.35) | 0.90 |  |
| **Total protein, g/day** |  |  |  |  |
| HR (95% CI)^a^ |  |  |  | 0.19 |
| <median | 347/2,404 | 1.24 (1.01-1.51) | 0.04 |  |
| ≥median | 328/2,403 | 1.26 (1.04-1.53) | 0.02 |  |
| **eGFR, mL/min/1.73 m2** |  |  |  |  |
| HR (95% CI)^a^ |  |  |  | 0.42 |
| <35 | 74/208 | 4.44 (1.94-10.16) | <0.001 |  |
| ≥35 | 601/4,599 | 1.21 (1.04-1.40) | 0.01 |  |
| **PRS (eGFR)** |  |  |  |  |
| HR (95% CI)^a^ |  |  |  | 0.93 |
| Low | 182/1,326 | 1.57 (1.18-2.08) | 0.002 |  |
| Medium | 195/1,326 | 1.41 (1.05-1.89) | 0.02 |  |
| High | 187/1,326 | 1.05 (0.79-1.38) | 0.76 |  |
| **CKD stage** |  |  |  |  |
| HR (95% CI)^a^ |  |  |  | 0.10 |
| 1-2 | 124/1,063 | 1.60 (1.13-2.27) | 0.008 |  |
| 3a-3b | 365/3,071 | 1.01 (0.83-1.23) | 0.93 |  |
| 4-5 | 144/515 | 1.30 (0.91-1.88) | 0.15 |  |
| ^a^Hazard Ratios with 95% Confidence Intervals (CI), adjusted for sex (excluding subgroup analysis), BMI, waist circumference, ethnicity, physical activity, smoking status (excluding subgroup analysis), education, alcohol intake (excluding subgroup analysis), energy intake, multimorbidity index, polypharmacy, Townsend deprivation index, prevalent diabetes, total protein intake (excluding subgroup analysis) and number of completed dietary assessments; stratified by age (5-year categories) (excluding subgroup analysis) and region.  Heterogeneity was tested by comparing two models – one without an interaction term between subgroup of interest and hPDI (categorical), with a model that included an interaction term. The likelihood ratio test was used to produce P-interaction values.  *Abbreviations: Q, quartile; uPDI, unhealthful plant-based diet index; BMI, Body Mass Index; HR, hazard ratio; CI, confidence interval; eGFR, estimated glomerular filtration rate; PRS, polygenic risk score; CKD, chronic kidney disease.* | | | | |

| **Table S10.** Sensitivity analyses showing hazard ratios (95% confidence intervals) across sex-specific healthful vs unhealthful plant-based diet index quartiles (Q), further adjusting for eGFR for participants who completed 1 or more dietary assessments and the associated risk of mortality (n=4,698) | | | | | |
| --- | --- | --- | --- | --- | --- |
| **hPDI** | Q1 | Q2 | Q3 | Q4 | P-trend |
| Cases/total | 225/1,265 | 166/1,217 | 137/1,109 | 128/1,107 |  |
| HR (95% CI)^a^ | 1.00^b^ | 0.81 (0.66-1.00) | 0.74 (0.59-0.92) | 0.68 (0.54-0.86) | <0.001 |
| **uPDI** | Q1 | Q2 | Q3 | Q4 | P-trend |
| Cases/total | 145/1,226 | 174/1,353 | 155/1,222 | 182/997 |  |
| HR (95% CI)^a^ | 1.00^b^ | 1.08 (0.86-1.35) | 1.23 (0.97-1.56) | 1.46 (1.15- 1.86) | 0.009 |
| ^a^Hazard Ratios with 95% Confidence Intervals (CI), adjusted for sex, BMI, waist circumference, ethnicity, physical activity, smoking status, education, alcohol intake, energy intake, multimorbidity index, polypharmacy, Townsend deprivation index, prevalent diabetes, total protein intake, number of completed dietary assessments and eGFR; stratified by age (5-year categories) and region.  P-trend is for linear trend.  ^b^Reference categories.  *Abbreviations: Q, quartile; hPDI, healthful plant-based diet index; uPDI, unhealthful plant-based diet index; BMI, Body Mass Index; eGFR, estimated glomerular filtration rate; HR, hazard ratio; CI, confidence interval.* | | | | | |

| **Table S11.** Sensitivity analyses showing hazard ratios (95% confidence intervals) across sex-specific healthful vs unhealthful plant-based diet index quartiles (Q), further adjusting for genetic susceptibility of kidney diseases, for participants who completed 1 or more dietary assessments and the associated risk of mortality (n=4,807) | | | | | |
| --- | --- | --- | --- | --- | --- |
| **hPDI** | Q1 | Q2 | Q3 | Q4 | P-trend |
| Cases/total | 236/1,309 | 168/1,239 | 140/1,131 | 131/1,128 |  |
| HR (95% CI)^a^ | 1.00^b^ | 0.78 (0.64-0.96) | 0.73 (0.58-0.91) | 0.67 (0.53-0.84) | <0.001 |
| **uPDI** | Q1 | Q2 | Q3 | Q4 | P-trend |
| Cases/total | 151/1,250 | 177/1,374 | 157/1,148 | 190/1,035 |  |
| HR (95% CI)^a^ | 1.00^b^ | 1.07 (0.86-1.34) | 1.22 (0.96-1.55) | 1.50 (1.18- 1.90) | 0.004 |
| ^a^Hazard Ratios with 95% Confidence Intervals (CI), adjusted for sex, BMI, waist circumference, ethnicity, physical activity, smoking status, education, alcohol intake, energy intake, multimorbidity index, polypharmacy, Townsend deprivation index, prevalent diabetes, total protein intake, number of completed dietary assessments and PRS (eGFR); stratified by age (5-year categories) and region.  P-trend is for linear trend.  ^b^Reference categories.  *Abbreviations: Q, quartile; hPDI, healthful plant-based diet index; uPDI, unhealthful plant-based diet index; BMI, Body Mass Index; eGFR, estimated glomerular filtration rate; HR, hazard ratio; CI, confidence interval.* | | | | | |

| **Table S12.** Sensitivity analyses showing hazard ratios (95% confidence intervals) across sex-specific healthful vs unhealthful plant-based diet index quartiles (Q), removing the first 2 years of follow-up for participants who completed 1 or more dietary assessments and the associated risk of mortality (n=4,700) | | | | | |
| --- | --- | --- | --- | --- | --- |
| **hPDI** | Q1 | Q2 | Q3 | Q4 | P-trend |
| Cases/total | 195/1,267 | 143/1,214 | 120/1,108 | 114/1,111 |  |
| HR (95% CI) |  |  |  |  |  |
| Model 1 | 1.00^a^ | 0.74 (0.59-0.92) | 0.66 (0.53-0.83) | 0.64 (0.51-0.81) | <0.001 |
| Model 2 | 1.00^a^ | 0.83 (0.66-1.03) | 0.76 (0.60-0.97) | 0.74 (0.57-0.94) | 0.009 |
| **uPDI** | Q1 | Q2 | Q3 | Q4 | P-trend |
| Cases/total | 124/1,221 | 156/1,353 | 133/1,122 | 159/1,004 |  |
| HR (95% CI) |  |  |  |  |  |
| Model 1 | 1.00^a^ | 1.18 (0.93-1.50) | 1.23 (0.96-1.57) | 1.72 (1.35-2.18) | <0.001 |
| Model 2 | 1.00^a^ | 1.17 (0.92-1.49) | 1.29 (1.00-1.67) | 1.59 (1.23- 2.07) | 0.007 |
| Model 1 adjusted for sex and education; stratified by age (5-year categories) and region.  Model 2: Model 1 plus BMI, waist circumference, ethnicity, physical activity, smoking status, alcohol intake, energy intake, multimorbidity index, polypharmacy, Townsend deprivation index, prevalent diabetes, total protein intake and number of completed dietary assessments.  P-trend is for linear trend.  ^a^Reference categories.  *Abbreviations: Q, quartile; hPDI, healthful plant-based diet index; uPDI, unhealthful plant-based diet index; BMI, Body Mass Index; HR, hazard ratio; CI, confidence interval.* | | | | | |

| **Table S13.** Sensitivity analyses showing hazard ratios (95% confidence intervals) across sex-specific healthful vs unhealthful plant-based diet index quartiles (Q), for participants who completed 2 or more dietary assessments and the associated risk of mortality (n=2,689) | | | | | |
| --- | --- | --- | --- | --- | --- |
| **hPDI** | Q1 | Q2 | Q3 | Q4 | P-trend |
| Cases/total | 134/806 | 82/751 | 69/608 | 49/524 |  |
| HR (95% CI) |  |  |  |  |  |
| Model 1 | 1.00^a^ | 0.60 (0.45-0.79) | 0.60 (0.45-0.81) | 0.55 (0.39-0.76) | <0.001 |
| Model 2 | 1.00^a^ | 0.69 (0.51-0.92) | 0.71 (0.52-0.97) | 0.62 (0.44-0.88) | 0.004 |
| **uPDI** | Q1 | Q2 | Q3 | Q4 | P-trend |
| Cases/total | 83/817 | 105/817 | 69/608 | 77/447 |  |
| HR (95% CI) |  |  |  |  |  |
| Model 1 | 1.00^a^ | 1.28 (0.95-1.71) | 1.14 (0.82-1.57) | 1.91 (1.39-2.63) | 0.001 |
| Model 2 | 1.00^a^ | 1.14 (0.84-1.55) | 1.02 (0.72-1.44) | 1.51 (1.06- 2.14) | 0.08 |
| Model 1 adjusted for sex and education; stratified by age (5-year categories) and region.  Model 2: Model 1 plus BMI, waist circumference, ethnicity, physical activity, smoking status, alcohol intake, energy intake, multimorbidity index, polypharmacy, Townsend deprivation index, prevalent diabetes, total protein intake and number of completed dietary assessments.  P-trend is for linear trend.  ^a^Reference categories.  *Abbreviations: Q, quartile; hPDI, healthful plant-based diet index; uPDI, unhealthful plant-based diet index; BMI, Body Mass Index; HR, hazard ratio; CI, confidence interval.* | | | | | |
